# Protecting Emergency Medical Services (EMS) Staff from Aggression and Violence in Conflict Encounters (PEACE1): A survey of Wales Adults attitudes in 2022

**DOI:** 10.1101/2023.11.30.23299241

**Authors:** Nigel Rees, Daniel Todd, Francesca Fiorentino, Peter O’Meara, Lauren Williams, Julia Williams, Claire Hawkes

## Abstract

**Background:** Emergency Medical Services (EMS) staff frequently encounter violence and aggression (V&A) directed towards them, which negatively affects their wellbeing and provision of services. This is an international problem and employers and others are tackling it through policy, education, legislation, and public information campaigns. The aim of this study was to explore the Welsh public’s views of V&A directed at EMS staff and their awareness of policy changes and the reach of media campaigns associated with recent policy changes.

**Methods:** In August 2022, we conducted a survey including a YouGov non probabilistic purposive sample of 1,010 Wales adults (aged 18+) from a matched panel representative of the population derived from a UK YouGov panel of >360,000 adults registered.

**Results:** Our sample included 52.4% women, median age and IQR was 52 and 52.5% were in social grade ABC1. 60.2% lived in areas with the most urban conurbations and 39.8% in the most rural conurbations. Most (62.5%) participants had heard of an instance of V&A directed towards EMS staff, but 24.4% had never heard, and 72.1% had never witnessed V&A. 15.5% had witnessed V&A directed towards EMS staff less than 10 times and these were more likely to be younger. 17.7% heard via work, and younger participants were more likely hear via social settings. 81.1% heard through media, and older participants were more likely to have heard through the media. 90.4% disagreed that V&A towards EMS staff can be acceptable in some cases and 53.3% were not aware of any related publicity and/or media campaigns. 92.4% thought intoxication with alcohol would likely contribute to V&A towards EMS staff, 90.5% thought intoxication with drugs and 84.3% thought an altered mental status following illness and/or injury would also likely contribute. 22% were aware of the Assaults on Emergency Workers Act, and 75.2% thought those intoxicated with drugs or alcohol would unlikely be deterred from V&A towards EMS staff by this act. 75.6% thought those with altered mental status following illness and/or injury would unlikely be deterred, and 42.2% thought it unlikely other members of the public would be deterred; 42.9% thought it was likely. Younger participants and those with a social grade of C2/D/E were more likely to think those intoxicated with drugs and alcohol, altered mental status following illness and/or injury and other members of the public would be deterred.

**Conclusion:** Understanding public attitudes towards V&A directed to EMS staff is important for their wellbeing and maintaining services. There appears to be good awareness of this issue in the Wales public, who also find V&A directed towards EMS staff unacceptable but underestimate the scale of the problem. Whilst the media appears to be the best form of communication, we did not find evidence of impact of current efforts to tackle this issues, as participants were largely unaware of legislation and information campaigns aimed at V&A directed towards EMS staff. Participants overwhelmingly felt current legislation would not deter V&A towards EMS staff, but younger people were more likely to think it would. We therefore recommend massages may be amplified, and targeted towards young people, men, those in social grade of C2/D/E and through social settings where V&A may be encountered more often and who were more likely to feel current legislation would deter. It is unclear if current efforts of policy, legislation and communication campaigns have any impact on talking V&A directed towards EMS staff, and we therefore recommend further research to understand and develop evidence-based interventions, along with tackling the influence of those factors involved, such as intoxication with drugs, alcohol, and altered mental status following illness and/or injury, by improving education, support, and care provision for these groups of people.

## Background

Emergency Medical Services (EMS) staff worldwide have long been at risk of encountering violence and aggression (V&A) directed towards them during their work (Copeland et al 2017, Yan et al 2013, Murray et al 2019, Rees & Whitfield 2005). For this study define EMS staff as Emergency Medical Technicians (EMT’s), Paramedics, Nurses, call handlers and others who provide prehospital and emergency care. V&A has been reported in 8.5% of patient encounters with USA EMS staff; 52.7% directed towards staff (Grange and Corbett 2002), and verbal forms of V&A are most prevalent (Bigham et al 2014, Oliver & Levine 2015, Bernaldo-de-Quiros et al 2014, Baydin et al 2014), along with physical injury, and on rare occasions, deaths occur (Maguire et al 2005).

Between 61% and 90% of EMS staff have reported being subjected to physical violence during their duties and 17% have reported being threatened with a weapon (Pozzi 1998, Corbett et al 1998, Suserud et al. 2002, Boyle et al 2007, Murray et al 2019). Exposure to such V&A can result in increased levels of stress, fear, anxiety, emotional exhaustion, and burnout syndrome (Taylor et al 2016, Yoon et al 2016, Bernaldo-De-Quirós et al 2015). This issue has been reported in the literature since 1978, but despite its significance, nearly half a century later, little effective progress has been made to tackle it (Murray et al 2019).

Many EMS systems have introduced interventions attempting to tackle this problem, and it has increasingly become a priority within Wales (UK), resulting in policy and legislation developments and initiatives. We have previously described the multi-agency approach taken in Wales (UK) (Rees et al 2021), which includes changes in legislation such as the Assaults on Emergency Workers (Offences) Act (2018), the Obligatory responses to violence in healthcare (2018) and the JESG #WithUsNotAgainst Us campaign. These approaches reflect the complexity of V&A directed towards EMS staff and acknowledges the need for further research and evidence-based interventions to better understand and effectively tackle this issue. This study (PEACE 1) is part of a program of Research and Innovation which aims to explore strategies designed to protect EMS staff from aggression and violence in conflict encounters.

The overall aim of this project was to explore the Welsh public’s views of V&A directed at EMS Staff and their awareness of policy changes and the reach of media campaigns associated with recent policy changes. The specific objectives were to:

- Explore general views on V&A directed towards EMS staff.
- Explore views on characteristics associated with V&A such as intoxication, drugs, altered mental status and the role of medical illnesses and mental health problems.
- Explore the impact of policy changes such as The Assaults on Emergency Workers (Offences) Act 2018 and #WithUsNotAgainst Us campaign in order to gauge the attitudes, understandings and impact of these initiatives in the public.

## Methods

### Survey design

Questions for the survey were developed from a previously published literature review by our team (Rees et al 2021), along with correspondence and discussion with people and groups with expertise in this area. We worked with YouGov and researchers familiar with this method (Hawkes et al 2017) to optimize question clarity of meaning and ease of understanding.

### Sample

We included a YouGov non probabilistic purposive sample of 1,010 Wales adults (aged 18+) from a matched panel representative of the population derived from a UK YouGov panel of >360,000 adults registered. The achieved sample was weighted to be representative of Wales adults (aged 18+) in terms of age, sex, social class, and type of newspaper chosen (YouGov 2022).

### Analysis

To assess the extent to which our study cohort was representative of the overall population, we expressed all participant characteristics as numbers, percentages, medians, and quartiles as appropriate. Where responses to questions were less than 10, they were grouped and reported with an appropriate category. The National Readership Survey classification system (2022) was used to categorise social grade (A: workers in high managerial, administrative, or professional jobs; B: workers in intermediate managerial, administrative, or professional jobs; C1: workers in supervisory, clerical, and junior managerial, administrative, or professional jobs; C2: skilled manual workers; D: semi- and unskilled manual workers; E: state pensioners, casual or lowest grade workers, and those unemployed with state benefits only).

We analysed survey response data using Stata V17.0 SE according to a predefined statistical analysis plan, using generalised linear regression models to obtain adjusted comparisons. The precise form of the model used reflected the nature of the variable under consideration (logistic models for binary variables; ordinal logistic models for ordered variables). We adjusted the models for prespecified variables which were judged to impact perceptions of EMS staff: gender, age, social grade, Welsh region and working status (working full or part time vs full time student, retired, unemployed or not working/other). All analysis was unweighted, consistent with our previous work using YouGov data (Hawkes et al 2017). Responses of ‘don’t know’ and ‘don’t know/can’t recall’ were excluded from ordinal regressions analysis due to not being unordered responses.

Responses to how many times have you witnessed violence and/or aggression directed towards EMS staff were dichotomised into No vs Yes, they had witnessed V&A. A sensitivity analysis explored which characteristics were more likely to be indicators of having witnessed violence and/or aggression directed towards EMS staff. Participants who had never heard about V&A incident/s were grouped with participants who reported having not heard about V&A incidents via a specific situation. A sensitivity analysis explored the implication of including these participants on characteristics more likely to have heard about V&A incident/s directed towards EMS staff via these situations.

### Ethical considerations

The study was approved by the Health Research Authority and Health and Care Research Wales (IRAS ref. 313346) and complied with Health Research Authority guidance (HRA, 2019). Participants were not patients, and the data set were pseudonymised. Organisational approval was received from the Medical and Clinical Services Directorate of the Welsh Ambulance Service NHS Trust

## Results

The online survey was conducted between 18th - 24th August 2022 and included a sample of 1,010 Wales adults (aged 18+). The characteristics of this sample are presented in table 1. 52.4% were women, and the median age and IQR was 52 [37,67], table 1. Just over half were in social grade ABC1 (52.5%). 60.2% lived in areas with the most urban conurbations and 39.8% in the areas that are most rural.

**Table 1:** Study cohort’s characteristics.

| Cohort's characteristics | Frequency (percentage) |
| --- | --- |
| Gender |  |
| Female | 529 (52.4%) |
| Male | 481 (47.6%) |
| Age (years) |  |
| p50 [p25, p75] | 52 [37, 67] |
| Social Grade |  |
| A/B/C1 | 530 (52.5%) |
| C2/D/E | 380 (47.5%) |
| Welsh region |  |
| Mid and West Wales | 193 (19.1%) |
| North Wales | 209 (20.7%) |
| Cardiff | 133 (13.2%) |
| South Wales Central | 69 (6.8%) |
| South Wales East | 235 (23.3%) |
| South Wales West | 171 (16.9%) |
| Working Status |  |
| No | 524 (51.9%) |
| ALL WORKERS | 486 (48.1%) |

### Experience of V&A directed towards EMS staff

When considering public experiences, 24.4% had never heard about V&A directed towards EMS staff, but 62.5% had heard of an instance, table 2. No characteristics were indicative of if a respondent was more likely to have heard about more incidents of V&A towards EMS staff, table 3. Most (72.1%) had never witnessed V&A directed towards EMS staff but 15.5% had done so less than 10 times, table 2.

**Table 2:**
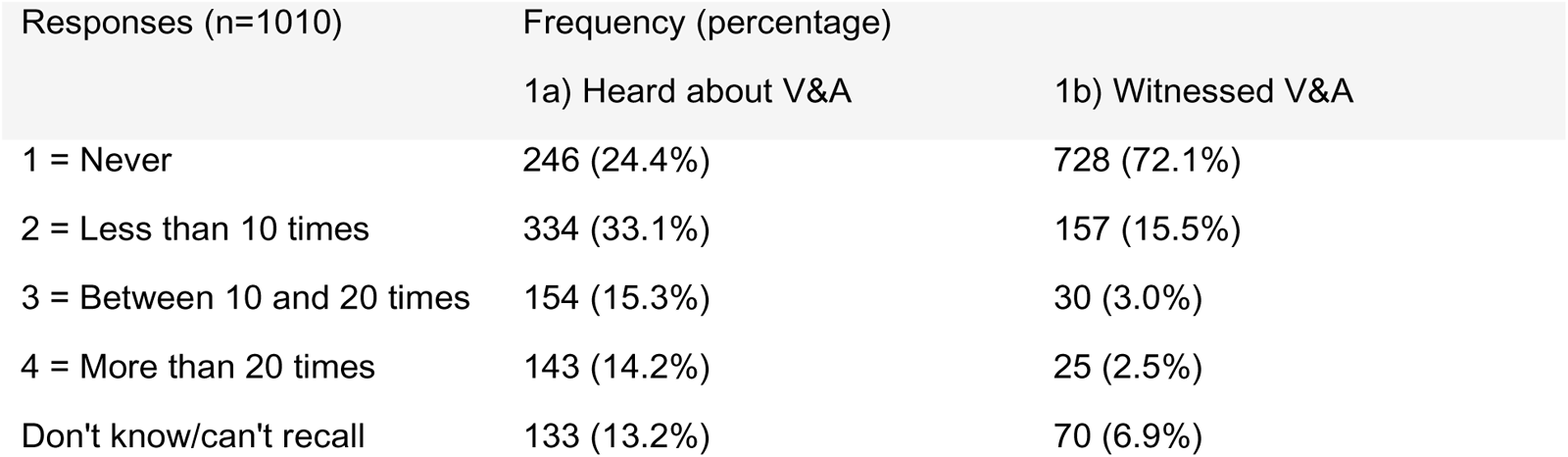
Tabulation of “How many times have you heard about or witnessed violence and/or aggression directed towards EMS staff?”

**Table 3:** Ordinal logistic regression of “How many times have you heard about violence and/or aggression directed towards EMS staff?”

| Q1a*) Ordinal logistic regression, Heard about V&A (n=877) | Univariate logistic regressions Odds Ratio, [95% C.I. of OR], (P-value) | Multivariable logistic regressions Odds Ratio, [95% C.I. of OR], (P-value) |
| --- | --- | --- |
| Female | 0.886 [0.696, 1.127] (p=0.324) | 0.878 [0.689, 1.119] (p=0.293) |
| Age | 0.994 [0.987, 1.000] (p=0.064) | 0.994 [0.986, 1.001] (p=0.094) |
| C2/D/E | 0.850 [0.668, 1.082] (p=0.187) | 0.846 [0.663, 1.080] (p=0.181) |
| Welsh region (Reference category = Cardiff) |  |  |
| Mid and West Wales | 0.933 [0.606, 1.438] (p=0.754) | 0.987 [0.638, 1.528] (p=0.954) |
| North Wales | 0.986 [0.648, 1.500] (p=0.948) | 1.021 [0.671, 1.555] (p=0.922) |
| South Wales Central | 0.844 [0.476, 1.496] (p=0.561) | 0.895 [0.503, 1.592] (p=0.706) |
| South Wales East | 1.123 [0.745, 1.691] (p=0.580) | 1.145 [0.760, 1.725] (p=0.516) |
| South Wales West | 0.882 [0.556, 1.399] (p=0.593) | 0.904 [0.570, 1.434] (p=0.668) |
| ALL WORKERS | 1.131 [0.890, 1.438] (p=0.314) | 0.992 [0.761, 1.295] (p=0.955) |

Note: \* The category 'don't know/can't recall' has been excluded from the model as the response is not ordered.

Younger participants [OR 0.972, 95% CI 0.962-0.981; p<0.001] were more likely to have witnessed more incidents of V&A directed towards EMS staff, but this included a low number of responses in the 10-20 and 20+ times categories, table 4. A sensitivity analysis explored grouping the categories into No vs Yes, they had witnessed V&A directed towards EMS staff. This also concluded that younger participants were more likely to have witness V&A incident/s [OR 0.973, 95% CI 0.964-0.983; p<0.001], table 5.

**Table 4:** Ordinal logistic regression of “How many times have you witnessed violence and/or aggression directed towards EMS staff?”

| Q1b*) Ordinal logistic regression, Witnessed V&A (n=940) | Univariate logistic regressions Odds Ratio, [95% C.I. of OR], (P-value) | Multivariable logistic regressions Odds Ratio, [95% C.I. of OR], (P-value) |
| --- | --- | --- |
| Female | 1.080 [0.796, 1.465] (p=0.621) | 1.035 [0.757, 1.416] (p=0.827) |
| Age | 0.974 [0.965, 0.983] (p<0.001) | 0.972 [0.962, 0.981] (p<0.001) |
| C2/D/E | 1.069 [0.788, 1.449] (p=0.668) | 1.042 [0.761, 1.427] (p=0.798) |
| Welsh region (Reference category = Cardiff) |  |  |
| Mid and West Wales | 1.027 [0.576, 1.831] (p=0.927) | 1.357 [0.748, 2.462] (p=0.315) |
| North Wales | 1.345 [0.777, 2.329] (p=0.289) | 1.546 [0.882, 2.710] (p=0.128) |
| South Wales Central | 0.959 [0.437, 2.106] (p=0.917) | 1.096 [0.493, 2.440] (p=0.822) |
| South Wales East | 1.433 [0.839, 2.448] (p=0.188) | 1.507 [0.874, 2.597] (p=0.140) |
| South Wales West | 1.564 [0.888, 2.754] (p=0.122) | 1.698 [0.953, 3.025] (p=0.072) |
| ALL WORKERS | 1.191 [0.878, 1.614] (p=0.261) | 0.824 [0.589, 1.153] (p=0.258) |

Note: \* The category 'don't know/can't recall' has been excluded from the model as the response is not ordered.

**Table 5:** Binary logistic regression of “No vs Yes, have you witnessed violence and/or aggression directed towards EMS staff?”

| Q1b*) Ordinal logistic regression, Witnessed V&A (n=940) | Univariate logistic regressions Odds Ratio, [95% C.I. of OR], (P-value) | Multivariable logistic regressions Odds Ratio, [95% C.I. of OR], (P-value) |
| --- | --- | --- |
| Female | 1.104 [0.812, 1.500] (p=0.528) | 1.067 [0.778, 1.463] (p=0.688) |
| Age | 0.974 [0.966, 0.983] (p<0.001) | 0.973 [0.964, 0.983] (p<0.001) |
| C2/D/E | 1.058 [0.779, 1.437] (p=0.717) | 1.042 [0.759, 1.430] (p=0.800) |
| Welsh region (Reference category = Cardiff) |  |  |
| Mid and West Wales | 0.970 [0.543, 1.732] (p=0.917) | 1.259 [0.692, 2.291] (p=0.451) |
| North Wales | 1.350 [0.777, 2.345] (p=0.288) | 1.550 [0.880, 2.730] (p=0.129) |
| South Wales Central | 0.917 [0.417, 2.017] (p=0.829) | 1.031 [0.461, 2.304] (p=0.941) |
| South Wales East | 1.417 [0.826, 2.429] (p=0.206) | 1.479 [0.854, 2.563] (p=0.162) |
| South Wales West | 1.542 [0.872, 2.728] (p=0.137) | 1.667 [0.931, 2.985] (p=0.086) |
| ALL WORKERS | 1.213 [0.893, 1.648] (p=0.216) | 0.861 [0.615, 1.206] (p=0.385) |

Note: \* The category 'don't know/can't recall' has been excluded from the model as the response is not ordered.

Of those who reported having heard about violence and/or aggressive directed towards EMS staff, 17.7% reported they had heard via work, table 6. Women [OR 1.648, 95% CI 1.065-2.548; p=0.025], younger participants [OR 0.982, 95% CI 0.968-0.996; p=0.010] and currently employed [OR 2.992, 95% CI 1.841-4.863; p<0.001] participants were more likely to have heard about violent and/or aggressive incident/s via work, table 7. The sensitivity analysis, including participants who had never heard about a V&A incident/s, also conclude the same characteristics were more likely to have heard about V&A incident/s via work, table 8.

**Table 6:** Tabulation of “In which of the following situations have you ever heard about violent and/or aggressive incident/s?”

| Q2) In question 1a, you reported having heard about violence and/or aggression towards EMS staff. In which of the following situations have you ever heard about violent and/or aggressive incident/s? (n=631) | Frequency (percentage) |
| --- | --- |
| Via work | 112 (17.7%) |
| Via a social setting (e.g. pub, sports team, etc.) | 118 (18.7%) |
| Through the media (e.g. TV reports, a campaign on social media, etc.) | 512 (81.1%) |
| Through discussions with friends/ acquaintances/ family members | 293 (46.4%) |
| Other | 20 (3.2%) |
| Don't know/can't recall | 8 (1.3%) |

**Table 7:** Binary logistic regression of “Had you ever heard about violent and/or aggressive incident/s via work?”

| Q2a*) Binary logistic regression, via work (if reported hearing about violence in Q1a) (n=623) | Univariate logistic regressions Odds Ratio, [95% C.I. of OR], (P-value) | Multivariable logistic regressions Odds Ratio, [95% C.I. of OR], (P-value) |
| --- | --- | --- |
| Female | 1.457 [0.960, 2.211] (p=0.077) | 1.648 [1.065, 2.548] (p=0.025) |
| Age | 0.974 [0.963, 0.986] (p<0.001) | 0.982 [0.968, 0.996] (p=0.010) |
| C2/D/E | 0.766 [0.505, 1.163] (p=0.210) | 0.837 [0.542, 1.294] (p=0.424) |
| Welsh region (Reference category = Cardiff) |  |  |
| Mid and West Wales | 0.853 [0.405, 1.798] (p=0.676) | 1.245 [0.569, 2.724] (p=0.584) |
| North Wales | 0.872 [0.426, 1.787] (p=0.709) | 1.057 [0.502, 2.226] (p=0.884) |
| South Wales Central | 0.629 [0.211, 1.873] (p=0.405) | 0.902 [0.290, 2.805] (p=0.858) |
| South Wales East | 1.008 [0.507, 2.002] (p=0.983) | 1.108 [0.544, 2.255] (p=0.777) |
| South Wales West | 1.354 [0.638, 2.872] (p=0.430) | 1.543 [0.705, 3.378] (p=0.278) |
| ALL WORKERS | 3.609 [2.284, 5.703] (p<0.001) | 2.992 [1.841, 4.863] (p<0.001) |

Note: \* The category 'don't know/can't recall' has been excluded from the model as they were not certain if they had or had not heard about violence and/or aggression towards EMS staff via work.

**Table 8:** Binary logistic regression of “Had you ever heard about violent and/or aggressive incident/s via work?” including respondents who had never heard about V&A incident/s.

| Q2a*) Binary logistic regression, via work (n = 869) | Univariate logistic regressions Odds Ratio, [95% C.I. of OR], (P-value) | Multivariable logistic regressions Odds Ratio, [95% C.I. of OR], (P-value) |
| --- | --- | --- |
| Female | 1.485 [0.991, 2.227] (p=0.056) | 1.613 [1.061, 2.453] (p=0.025) |
| Age | 0.973 [0.962, 0.985] (p<0.001) | 0.981 [0.968, 0.995] (p=0.006) |
| C2/D/E | 0.737 [0.491, 1.105] (p=0.139) | 0.843 [0.554, 1.282] (p=0.424) |
| Welsh region (Reference category = Cardiff) |  |  |
| Mid and West Wales | 0.828 [0.402, 1.706] (p=0.608) | 1.177 [0.554, 2.504] (p=0.671) |
| North Wales | 0.916 [0.456, 1.840] (p=0.806) | 1.093 [0.532, 2.242] (p=0.809) |
| South Wales Central | 0.628 [0.216, 1.825] (p=0.393) | 0.769 [0.257, 2.300] (p=0.639) |
| South Wales East | 1.101 [0.565, 2.145] (p=0.777) | 1.151 [0.579, 2.287] (p=0.688) |
| South Wales West | 1.136 [0.552, 2.338] (p=0.729) | 1.282 [0.609, 2.698] (p=0.514) |
| ALL WORKERS | 3.586 [2.295, 5.604] (p<0.001) | 2.911 [1.814, 4.673] (p<0.001) |

Note: \* The category 'don't know/can't recall' has been excluded from the model as they were not certain if they had or had not heard about violence and/or aggression towards EMS staff via work.

**Table 9:** Binary logistic regression of “Had you ever heard about violent and/or aggressive incident/s via a social setting?”

| Q2b*) Binary logistic regression, Via a social setting (e.g. pub, sports team, etc.) (if reported hearing about violence in Q1a) (n=623) | Univariate logistic regressions Odds Ratio, [95% C.I. of OR], (P-value) | Multivariable logistic regressions Odds Ratio, [95% C.I. of OR], (P-value) |
| --- | --- | --- |
| Female | 0.931 [0.624, 1.391] (p=0.729) | 0.956 [0.632, 1.446] (p=0.831) |
| Age | 0.972 [0.960, 0.983] (p<0.001) | 0.972 [0.960, 0.985] (p<0.001) |
| C2/D/E | 1.151 [0.770, 1.721] (p=0.492) | 1.180 [0.779, 1.789] (p=0.435) |
| Welsh region (Reference category = Cardiff) |  |  |
| Mid and West Wales | 0.870 [0.405, 1.868] (p=0.721) | 1.160 [0.527, 2.556] (p=0.712) |
| North Wales | 1.317 [0.652, 2.662] (p=0.443) | 1.569 [0.761, 3.234] (p=0.222) |
| South Wales Central | 0.684 [0.228, 2.054] (p=0.498) | 0.822 [0.267, 2.534] (p=0.733) |
| South Wales East | 1.096 [0.545, 2.206] (p=0.797) | 1.177 [0.576, 2.404] (p=0.655) |
| South Wales West | 1.570 [0.736, 3.351] (p=0.243) | 1.811 [0.833, 3.937] (p=0.134) |
| ALL WORKERS | 1.695 [1.127, 2.548] (p=0.011) | 1.234 [0.796, 1.912] (p=0.347) |

Note: \* The category 'don't know/can't recall' has been excluded from the model as they were not certain if they had or had not heard about violence and/or aggression towards EMS staff via work.

Of those who reported having heard about violence and/or aggressive directed towards EMS staff, 18.7% reported they heard via a social setting, table 6. Younger participants [OR 0.972, 95% CI 0.960-0.985]; p<0.001] were more likely to have heard about violent and/or aggressive incident/s via a social setting, table XYZ. The sensitivity analysis including participants who had never heard about a V&A incident/s also conclude the same characteristics were more likely to have heard about V&A incident/s via a social setting, table 10.

**Table 10:** Binary logistic regression of “Had you ever heard about violent and/or aggressive incident/s via a social setting?” including respondents who had never heard about V&A incident/s.

| Q2b*) Binary logistic regression, Via a social setting (e.g. pub, sports team, etc.) (n=869) | Univariate logistic regressions Odds Ratio, [95% C.I. of OR], (P-value) | Multivariable logistic regressions Odds Ratio, [95% C.I. of OR], (P-value) |
| --- | --- | --- |
| Female | 0.975 [0.661, 1.438] (p=0.898) | 0.983 [0.660, 1.465] (p=0.934) |
| Age | 0.971 [0.959, 0.982] (p<0.001) | 0.972 [0.960, 0.984] (p<0.001) |
| C2/D/E | 1.080 [0.732, 1.594] (p=0.697) | 1.120 [0.750, 1.674] (p=0.579) |
| Welsh region (Reference category = Cardiff) |  |  |
| Mid and West Wales | 0.843 [0.401, 1.772] (p=0.652) | 1.148 [0.534, 2.466] (p=0.724) |
| North Wales | 1.351 [0.683, 2.674] (p=0.387) | 1.573 [0.783, 3.161] (p=0.203) |
| South Wales Central | 0.680 [0.232, 1.992] (p=0.482) | 0.807 [0.271, 2.403] (p=0.700) |
| South Wales East | 1.192 [0.604, 2.353] (p=0.613) | 1.276 [0.639, 2.550] (p=0.490) |
| South Wales West | 1.303 [0.629, 2.697] (p=0.477) | 1.442 [0.687, 3.026] (p=0.333) |
| ALL WORKERS | 1.749 [1.178, 2.598] (p=0.006) | 1.280 [0.839, 1.953] (p=0.252) |

Note: \* The category 'don't know/can't recall' has been excluded from the model as they were not certain if they had or had not heard about violence and/or aggression towards EMS staff via work.

In those who reported having heard about violence and/or aggressive directed towards EMS staff, 81.1% reported they heard through the media, table 6. Older participants [OR 1.016, 95% CI 1.002-1.029; p=0.021] were more likely to have heard about violent and/or aggressive incident/s through the media, table 11. Whilst participants from North Wales [OR 0.449, 95% CI 0.210-0.962; p=0.039] were less likely to have heard about violent and/or aggressive incidents than participants from Cardiff through the media, table 11. However, the sensitivity analysis, including participants who had never heard about a V&A incident/s, concluded that no characteristics were indicative of if a person was more likely to have heard about violent and/or aggressive incident/s directed towards EMS staff through the media, table 12.

**Table 11:** Binary logistic regression of “Had you ever heard about violent and/or aggressive incident/s through media?”

| Q2c*) Binary logistic regression, Through the media (e.g. TV reports, a campaign on social media, etc.) (if reported hearing about violence in Q1a) (n=623) | Univariate logistic regressions Odds Ratio, [95% C.I. of OR], (P-value) | Multivariable logistic regressions Odds Ratio, [95% C.I. of OR], (P-value) |
| --- | --- | --- |
| Female | 0.699 [0.460, 1.062] (p=0.093) | 0.667 [0.435, 1.023] (p=0.064) |
| Age | 1.018 [1.006, 1.030] (p=0.004) | 1.016 [1.002, 1.029] (p=0.021) |
| C2/D/E | 1.119 [0.739, 1.695] (p=0.594) | 1.111 [0.726, 1.701] (p=0.626) |
| Welsh region (Reference category = Cardiff) |  |  |
| Mid and West Wales | 0.864 [0.384, 1.943] (p=0.724) | 0.661 [0.288, 1.515] (p=0.328) |
| North Wales | 0.525 [0.249, 1.111] (p=0.092) | 0.449 [0.210, 0.962] (p=0.039) |
| South Wales Central | 0.541 [0.204, 1.438] (p=0.218) | 0.437 [0.161, 1.190] (p=0.105) |
| South Wales East | 0.780 [0.365, 1.665] (p=0.521) | 0.727 [0.337, 1.565] (p=0.415) |
| South Wales West | 0.797 [0.339, 1.876] (p=0.603) | 0.728 [0.306, 1.731] (p=0.472) |
| ALL WORKERS | 0.589 [0.388, 0.894] (p=0.013) | 0.663 [0.421, 1.046] (p=0.077) |

Note: \* The category 'don't know/can't recall' has been excluded from the model as they were not certain if they had or had not heard about violence and/or aggression towards EMS staff via work.

**Table 12:** Binary logistic regression of “Had you ever heard about violent and/or aggressive incident/s through media?” including respondents who had never heard about V&A incident/s.

| Q2c*) Binary logistic regression, Through the media (e.g. TV reports, a campaign on social media, etc.) (n = 869) | Univariate logistic regressions Odds Ratio, [95% C.I. of OR], (P-value) | Multivariable logistic regressions Odds Ratio, [95% C.I. of OR], (P-value) |
| --- | --- | --- |
| Female | 0.936 [0.714, 1.226] (p=0.630) | 0.921 [0.701, 1.211] (p=0.556) |
| Age | 1.002 [0.994, 1.009] (p=0.684) | 1.001 [0.992, 1.009] (p=0.843) |
| C2/D/E | 0.935 [0.713, 1.227] (p=0.629) | 0.938 [0.713, 1.236] (p=0.651) |
| Welsh region (Reference category = Cardiff) |  |  |
| Mid and West Wales | 0.869 [0.533, 1.415] (p=0.572) | 0.847 [0.515, 1.390] (p=0.511) |
| North Wales | 0.822 [0.510, 1.325] (p=0.420) | 0.814 [0.504, 1.315] (p=0.400) |
| South Wales Central | 0.732 [0.384, 1.396] (p=0.344) | 0.727 [0.380, 1.390] (p=0.334) |
| South Wales East | 1.112 [0.692, 1.787] (p=0.660) | 1.114 [0.692, 1.791] (p=0.657) |
| South Wales West | 0.681 [0.410, 1.133] (p=0.139) | 0.679 [0.408, 1.131] (p=0.137) |
| ALL WORKERS | 0.911 [0.695, 1.195] (p=0.502) | 0.892 [0.662, 1.203] (p=0.455) |

Note: \* The category 'don't know/can't recall' has been excluded from the model as they were not certain if they had or had not heard about violence and/or aggression towards EMS staff via work.

Of those who reported having heard about violence and/or aggressive directed towards EMS staff, 46.4% reported they heard through discussion with friends/acquaintances/family members, table 6. Females [OR 1.754, 95% CI 1.266-2.428; p=0.001] and participants from South Wales, compared to Cardiff, [OR 2.138, 95% CI 1.138-4.014; p=0.018] were more likely to have heard through discussions, table 13. The sensitivity analysis, including participants who had never heard about a V&A incident/s, conclude that females [OR 1.626, 95% CI 1.218-2.171; p=0.001] and younger [OR 0.991, 95% CI 0.982-1.000; p=0.041] participants were more likely to have heard through discussions, table 14. However, participants from South Wales, compared to Cardiff, were now no longer more likely to have heard through discussion, table 14.

**Table 13:** Binary logistic regression of “Had you ever heard about violent and/or aggressive incident/s through discussions?”

| Q2d*) Binary logistic regression, Through discussions with friends/ acquaintances/ family members (if reported hearing about violence in Q1a) (n=623) | Univariate logistic regressions Odds Ratio, [95% C.I. of OR], (P-value) | Multivariable logistic regressions Odds Ratio, [95% C.I. of OR], (P-value) |
| --- | --- | --- |
| Female | 1.698 [1.235, 2.335] (p=0.001) | 1.754 [1.266, 2.428] (p=0.001) |
| Age | 0.991 [0.982, 0.999] (p=0.037) | 0.992 [0.982, 1.002] (p=0.104) |
| C2/D/E | 1.007 [0.734, 1.381] (p=0.967) | 1.018 [0.735, 1.411] (p=0.915) |
| Welsh region (Reference category = Cardiff) |  |  |
| Mid and West Wales | 0.951 [0.538, 1.681] (p=0.864) | 1.136 [0.632, 2.042] (p=0.671) |
| North Wales | 0.827 [0.476, 1.436] (p=0.499) | 0.897 [0.511, 1.574] (p=0.704) |
| South Wales Central | 0.973 [0.455, 2.083] (p=0.944) | 1.043 [0.479, 2.272] (p=0.915) |
| South Wales East | 1.063 [0.622, 1.817] (p=0.822) | 1.108 [0.643, 1.907] (p=0.712) |
| South Wales West | 1.970 [1.060, 3.659] (p=0.032) | 2.138 [1.138, 4.014] (p=0.018) |
| ALL WORKERS | 1.251 [0.913, 1.714] (p=0.164) | 1.153 [0.811, 1.641] (p=0.428) |

Note: \* The category 'don't know/can't recall' has been excluded from the model as they were not certain if they had or had not heard about violence and/or aggression towards EMS staff via work.

**Table 14:** Binary logistic regression of “Had you ever heard about violent and/or aggressive incident/s through discussions?” including respondents who had never heard about V&A incident/s.

| Q2d*) Binary logistic regression, Through discussions with friends/ acquaintances/ family members (n = 869) | Univariate logistic regressions Odds Ratio, [95% C.I. of OR], (P-value) | Multivariable logistic regressions Odds Ratio, [95% C.I. of OR], (P-value) |
| --- | --- | --- |
| Female | 1.612 [1.212, 2.145] (p=0.001) | 1.626 [1.218, 2.171] (p=0.001) |
| Age | 0.989 [0.981, 0.997] (p=0.006) | 0.991 [0.982, 1.000] (p=0.041) |
| C2/D/E | 0.936 [0.705, 1.241] (p=0.646) | 0.963 [0.721, 1.286] (p=0.796) |
| Welsh region (Reference category = Cardiff) |  |  |
| Mid and West Wales | 0.913 [0.548, 1.522] (p=0.727) | 1.076 [0.637, 1.818] (p=0.784) |
| North Wales | 0.901 [0.546, 1.487] (p=0.682) | 0.967 [0.582, 1.607] (p=0.897) |
| South Wales Central | 0.948 [0.479, 1.877] (p=0.878) | 1.006 [0.503, 2.011] (p=0.987) |
| South Wales East | 1.183 [0.729, 1.918] (p=0.497) | 1.220 [0.747, 1.990] (p=0.427) |
| South Wales West | 1.328 [0.787, 2.240] (p=0.288) | 1.407 [0.828, 2.392] (p=0.207) |
| ALL WORKERS | 1.300 [0.981, 1.724] (p=0.068) | 1.169 [0.856, 1.595] (p=0.325) |

Note: \* The category 'don't know/can't recall' has been excluded from the model as they were not certain if they had or had not heard about violence and/or aggression towards EMS staff via work.

### Acceptability of V&A directed towards EMS staff

90.4% of participants disagreed with the statement “V&A towards EMS staff can be acceptable in some cases” table 15. Younger [OR 0.966, 95% CI 0.956-0.977; p<0.001] and social grades of C2/D/E [OR 1.859, 95% CI 1.301-2.658; p=0.001] participants were more likely to agree with statement, table 16.

**Table 15:** Tabulation of “To what extent, do you agree or disagree with the following statement? “Violence and/or aggression towards EMS staff can be acceptable in some cases”.

| Q3) To what extent, do you agree or disagree with the following statement? "Violence and/ or aggression towards EMS staff can be acceptable in some cases" (n=1010) | Frequency (percentage) |
| --- | --- |
| 1 = Strongly disagree | 828 (82.0%) |
| 2 = Tend to disagree | 85 (8.4%) |
| 3 = Neither agree nor disagree | 34 (3.4%) |
| 4 = Strongly agree/Tend to agree | 41 (4.1%) |
| Don't know | 22 (2.2%) |

**Table 16:** Ordinal logistic regression of “To what extent, do you agree or disagree with the following statement? “Violence and/or aggression towards EMS staff can be acceptable in some cases”.

| Q3*) Ordinal logistic regression. To what extent, do you agree or disagree with the following statement?<br>"Violence and/ or aggression towards EMS staff can be acceptable in some cases" | Univariate logistic regressions Odds Ratio, [95% C.I. of OR], (P-value) | Multivariable logistic regressions Odds Ratio, [95% C.I. of OR], (P-value) |
| --- | --- | --- |
| Female | 1.220 [0.869, 1.713] (p=0.251) | 1.189 [0.836, 1.690] (p=0.335) |
| Age | 0.970 [0.960, 0.980] (p<0.001) | 0.966 [0.956, 0.977] (p<0.001) |
| C2/D/E | 1.842 [1.307, 2.596] (p<0.001) | 1.859 [1.301, 2.658] (p=0.001) |
| Welsh region (Reference category = Cardiff) |  |  |
| Mid and West Wales | 0.867 [0.455, 1.654] (p=0.665) | 1.115 [0.571, 2.177] (p=0.750) |
| North Wales | 1.367 [0.751, 2.485] (p=0.306) | 1.543 [0.833, 2.861] (p=0.168) |
| South Wales Central | 1.363 [0.631, 2.947] (p=0.431) | 1.372 [0.619, 3.038] (p=0.436) |
| South Wales East | 0.947 [0.514, 1.744] (p=0.861) | 0.963 [0.515, 1.800] (p=0.905) |
| South Wales West | 1.563 [0.847, 2.884] (p=0.153) | 1.675 [0.892, 3.143] (p=0.108) |
| All workers | 0.994 [0.710, 1.393] (p=0.974) | 0.755 [0.525, 1.087] (p=0.130) |

Note: \* The category 'don't know' has been excluded from the model as the response is not ordered.

### Perceptions on the scale of V&A directed towards EMS staff

Perceptions varied on how many incidents of V&A directed towards EMS staff were reported to police each year in Wales. 33.2% of participants thought there are more than 500 incidents reported to police each year in Wales whilst 27.7% did not know, table 17. Males [OR 0.729, 95% CI 0.556-0.956; p=0.022] were more likely to think there were more incidents of V&A towards EMS staff reported to the police each year, table 18).

**Table 17:** Tabulation of “Approximately, how many incidents of violence and/or aggression towards EMS staff do you think are reported to the police each year in Wales?”

| Q5) Approximately, how many incidents of violence and/or aggression towards EMS staff do you think are reported to the police each year in Wales? (n=1010) | Frequency (percentage) |
| --- | --- |
| 1 = None | 11 (1.1%) |
| 2 = Less than 100 | 57 (5.6%) |
| 3 = More than 100, less than 200 | 94 (9.3%) |
| 4 = More than 200, less than 300 | 86 (8.5%) |
| 5 = More than 300, less than 400 | 76 (7.5%) |
| 6 = More than 400, less than 500 | 71 (7.0%) |
| 7 = More than 500 | 335 (33.2%) |
| Don't know | 280 (27.7%) |

**Table 18:** Ordinal logistic regression “Approximately, how many incidents of violence and/or aggression towards EMS staff do you think are reported to the police each year in Wales?”

| Q5*) Ordinal logistic regression. Approximately, how many incidents of violence and/or aggression towards EMS staff do you think are reported to the police each year in Wales? (n=730) | Univariate logistic regressions Odds Ratio, [95% C.I. of OR], (P-value) | Multivariable logistic regressions Odds Ratio, [95% C.I. of OR], (P-value) |
| --- | --- | --- |
| Female | 0.725 [0.555, 0.948] (p=0.019) | 0.729 [0.556, 0.956] (p=0.022) |
| Age | 1.006 [0.999, 1.014] (p=0.109) | 1.007 [0.999, 1.015] (p=0.082) |
| C2/D/E | 0.902 [0.692, 1.178] (p=0.450) | 0.884 [0.674, 1.158] (p=0.371) |
| Welsh region (Reference category = Cardiff) |  |  |
| Mid and West Wales | 1.291 [0.807, 2.066] (p=0.286) | 1.206 [0.747, 1.947] (p=0.444) |
| North Wales | 1.001 [0.638, 1.571] (p=0.995) | 0.986 [0.626, 1.552] (p=0.951) |
| South Wales Central | 1.491 [0.794, 2.798] (p=0.214) | 1.558 [0.825, 2.942] (p=0.172) |
| South Wales East | 0.929 [0.599, 1.441] (p=0.744) | 0.922 [0.594, 1.432] (p=0.718) |
| South Wales West | 1.478 [0.909, 2.404] (p=0.116) | 1.493 [0.916, 2.433] (p=0.108) |
| All workers | 1.069 [0.820, 1.394] (p=0.624) | 1.159 [0.869, 1.544] (p=0.315) |

Note: \* The category 'don't know' has been excluded from the model as the response is not ordered.

We asked participants approximately what percentage of EMS staff members they thought either have or will experience being subjected to physical violence (e.g. spitting, punching, kicking etc.) during their duties at least once in their career? Around 61.6% of participants thought 30% or more EMS staff members have, or will experience, being subjected to physical violence with 15.0% responding they did not know, table 19. No characteristics were indicative of if a respondent was more likely to have thought EMS staff have, or will experience, being subjected to physical violence, table 20. 60.4% of participants thought 10% or more EMS staff members either have, or will experience, being threatened with a weapon with 17.1% responding they did not know, table 21. No characteristics were indicative of if a respondent was more likely to have thought EMS staff have, or will experience, being threatened with a weapon, table 22.

**Table 19:** Tabulation of “What percentage of currently employed EMS staff members do you think either have or will experience being subjected to physical violence (e.g. spitting, punching, kicking etc.) during their duties?”

| Q6a) What percentage of currently employed EMS staff members do you think either have or will experience being subjected to physical violence (e.g. spitting, punching, kicking etc.) during their duties? (n=1010) | Frequency (percentage) |
| --- | --- |
| 1 = None/Less than 10%** | 56 (5.5%) |
| 2 = More than 10%, less than 30% | 180 (17.8%) |
| 3 = More than 30%, less than 60% | 262 (25.9%) |
| 4 = More than 60%, less than 90% | 224 (22.2%) |
| 5 = More than 90% | 136 (13.5%) |
| Don't know | 152 (15.0%) |

Note: \*\* The category has been grouped with an appropriate category due to low numbers.

**Table 20:** Ordinal logistic regression of “What percentage of currently employed EMS staff members do you think either have or will experience being subjected to physical violence (e.g. spitting, punching, kicking etc.) during their duties?”

| Q6a*) Ordinal logistic regression. What percentage of currently employed EMS staff members do you think either have or will experience being subjected to physical violence (e.g. spitting, punching, kicking etc.) during their duties? (n=858) | Univariate logistic regressions Odds Ratio, [95% C.I. of OR], (P-value) | Multivariable logistic regressions Odds Ratio, [95% C.I. of OR], (P-value) |
| --- | --- | --- |
| Female | 1.112 [0.875, 1.415] (p=0.384) | 1.143 [0.896, 1.457] (p=0.281) |
| Age | 0.999 [0.992, 1.006] (p=0.783) | 1.001 [0.994, 1.009] (p=0.727) |
| C2/D/E | 1.027 [0.808, 1.305] (p=0.831) | 1.056 [0.828, 1.345] (p=0.661) |
| Welsh region (Reference category = Cardiff) |  |  |
| Mid and West Wales | 1.045 [0.677, 1.615] (p=0.842) | 1.068 [0.687, 1.661] (p=0.769) |
| North Wales | 0.953 [0.629, 1.445] (p=0.821) | 0.959 [0.632, 1.457] (p=0.845) |
| South Wales Central | 0.821 [0.465, 1.449] (p=0.496) | 0.831 [0.470, 1.470] (p=0.525) |
| South Wales East | 1.212 [0.808, 1.817] (p=0.352) | 1.206 [0.804, 1.809] (p=0.365) |
| South Wales West | 1.043 [0.677, 1.608] (p=0.847) | 1.041 [0.675, 1.606] (p=0.855) |
| All workers | 1.188 [0.935, 1.509] (p=0.159) | 1.218 [0.935, 1.586] (p=0.144) |

Note: \* The category 'don't know' has been excluded from the model as the response is not ordered.

**Table 21:** Tabulation of “What percentage of currently employed EMS staff members do you think either have or will experience being threatened with a weapon during their duties?”

| Q6b) What percentage of currently employed EMS staff members do you think either have or will experience being threatened with a weapon during their duties? (n=1010) | Frequency (percentage) |
| --- | --- |
| 1 = None | 13 (1.3%) |
| 2 = Less than 10% | 215 (21.3%) |
| 3 = More than 10%, less than 30% | 251 (24.9%) |
| 4 = More than 30%, less than 60% | 219 (21.7%) |
| 5 = More than 60%, less than 90% | 101 (10.0%) |
| 6 = More than 90% | 38 (3.8%) |
| Don't know | 173 (17.1%) |

**Table 22:** Ordinal logistic regression of “What percentage of currently employed EMS staff members do you think either have or will experience being threatened with a weapon during their duties?”

| Q6b*) Ordinal logistic regression. What percentage of currently employed EMS staff members do you think either have or will experience being threatened with a weapon during their duties? (n=837) | Univariate logistic regressions Odds Ratio, [95% C.I. of OR], (P-value) | Multivariable logistic regressions Odds Ratio, [95% C.I. of OR], (P-value) |
| --- | --- | --- |
| Female | 0.971 [0.762, 1.238] (p=0.813) | 0.971 [0.759, 1.241] (p=0.811) |
| Age | 0.995 [0.988, 1.002] (p=0.128) | 0.998 [0.990, 1.005] (p=0.540) |
| C2/D/E | 1.200 [0.941, 1.530] (p=0.143) | 1.227 [0.960, 1.568] (p=0.103) |
| Welsh region (Reference category = Cardiff) |  |  |
| Mid and West Wales | 0.817 [0.531, 1.259] (p=0.360) | 0.841 [0.543, 1.305] (p=0.441) |
| North Wales | 1.148 [0.756, 1.743] (p=0.517) | 1.170 [0.768, 1.781] (p=0.465) |
| South Wales Central | 1.292 [0.731, 2.282] (p=0.378) | 1.303 [0.735, 2.307] (p=0.365) |
| South Wales East | 1.292 [0.859, 1.943] (p=0.218) | 1.283 [0.852, 1.931] (p=0.233) |
| South Wales West | 1.178 [0.761, 1.823] (p=0.462) | 1.169 [0.755, 1.810] (p=0.484) |
| All workers | 1.276 [1.001, 1.628] (p=0.049) | 1.236 [0.947, 1.613] (p=0.119) |

Note: \* The category 'don't know' has been excluded from the model as the response is not ordered.

84.3% of participants thought it to be very, or fairly likely that an EMS member of staff would experience verbal aggression (e.g. intimidation, threatening words or behaviour, etc.) from members of the public while they are at work; only 5.0% thought it was not at all/very likely with 10.6% responding they did not know, table 23. Older [OR 1.009, 95% CI 1.001-1.018; p=0.035] participants were more likely to think EMS staff would experience verbal aggression while at work, table 24. Participants from South Wales Central, compared to Cardiff, [OR 0.420, 95% CI 0.225-0.782; p=0.006] were less likely to think EMS staff would experience verbal aggression while at work, table 24.

**Table 23:** Tabulation of “ How likely or unlikely do you think it is that an EMS member of staff would experience verbal aggression from members of the public while they are at work?”

| Q8a) How likely or unlikely do you think it is that an EMS member of staff would experience verbal aggression from members of the public while they are at work? (n=1010) | Frequency (percentage) |
| --- | --- |
| 1 = Not at all likely/ Not very likely | 51** (5.0%) |
| 2 = Fairly likely | 279 (27.6%) |
| 3 = Very likely | 573 (56.7%) |
| Don't know | 107 (10.6%) |

**Table 24:**
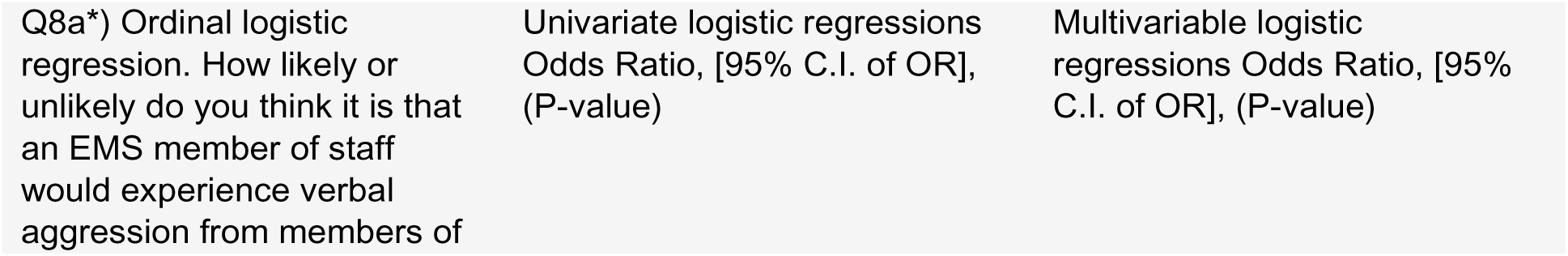
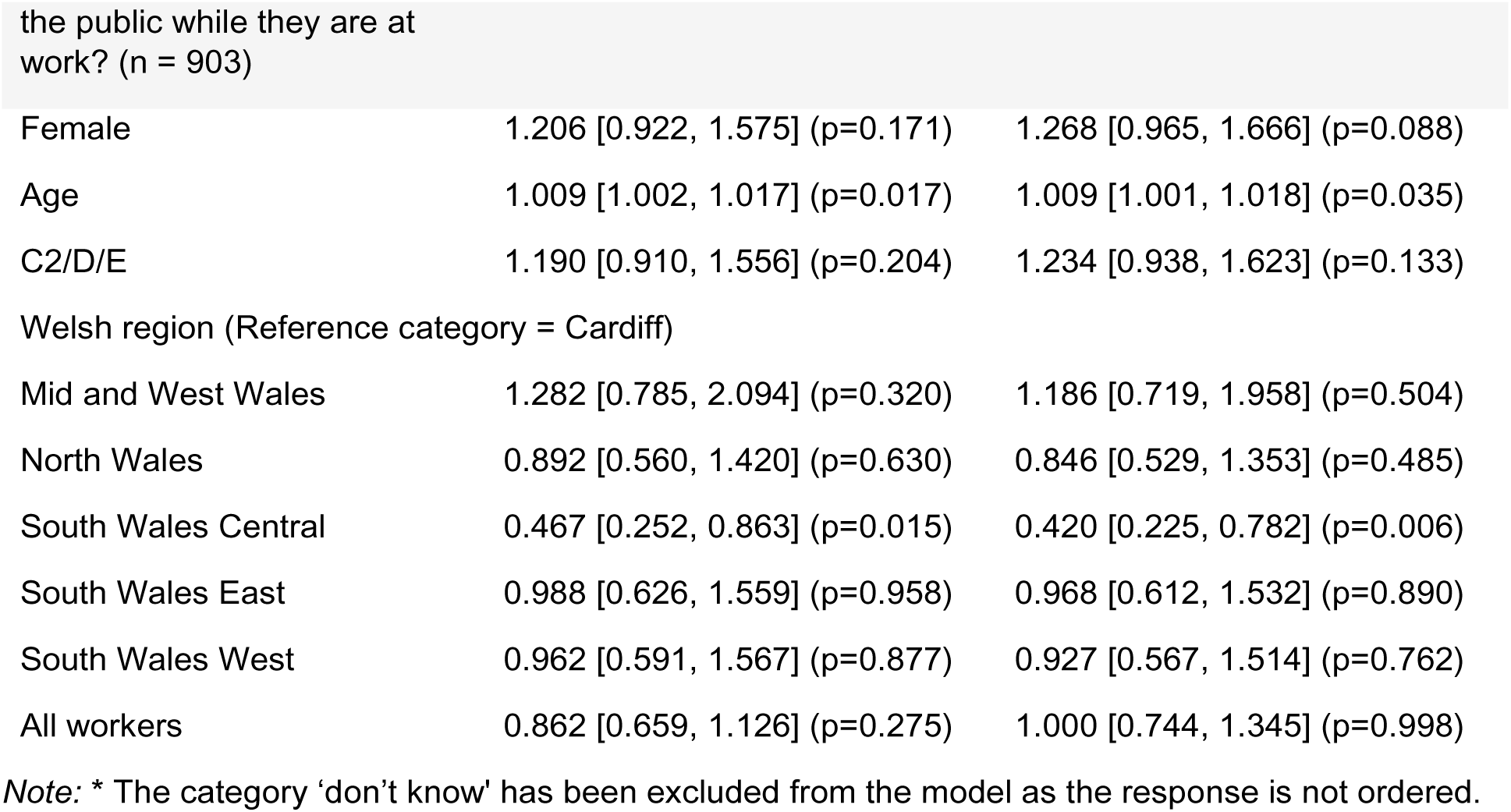
Ordinal logistic regression of “ How likely or unlikely do you think it is that an EMS member of staff would experience verbal aggression from members of the public while they are at work?”

| Q8a*) Ordinal logistic regression. How likely or unlikely do you think it is that an EMS member of staff would experience verbal aggression from members of | Univariate logistic regressions Odds Ratio, [95% C.I. of OR], (P-value) | Multivariable logistic regressions Odds Ratio, [95% C.I. of OR], (P-value) |
| --- | --- | --- |
| Female | 1.206 [0.922, 1.575] (p=0.171) | 1.268 [0.965, 1.666] (p=0.088) |
| Age | 1.009 [1.002, 1.017] (p=0.017) | 1.009 [1.001, 1.018] (p=0.035) |
| C2/D/E | 1.190 [0.910, 1.556] (p=0.204) | 1.234 [0.938, 1.623] (p=0.133) |
| Welsh region (Reference category = Cardiff) |  |  |
| Mid and West Wales | 1.282 [0.785, 2.094] (p=0.320) | 1.186 [0.719, 1.958] (p=0.504) |
| North Wales | 0.892 [0.560, 1.420] (p=0.630) | 0.846 [0.529, 1.353] (p=0.485) |
| South Wales Central | 0.467 [0.252, 0.863] (p=0.015) | 0.420 [0.225, 0.782] (p=0.006) |
| South Wales East | 0.988 [0.626, 1.559] (p=0.958) | 0.968 [0.612, 1.532] (p=0.890) |
| South Wales West | 0.962 [0.591, 1.567] (p=0.877) | 0.927 [0.567, 1.514] (p=0.762) |
| All workers | 0.862 [0.659, 1.126] (p=0.275) | 1.000 [0.744, 1.345] (p=0.998) |

Note: \* The category 'don't know' has been excluded from the model as the response is not ordered.

Most participants (71.9%) felt it was very, or fairly likely that an EMS member of staff would experience physical violence (e.g. spitting, punching, kicking, etc.) from members of the public while they are at work; 16.0% thought it was not at all/very likely with 12.0% responding they did not know, table 25. Females [OR 1.405, 95% CI 1.089-1.813; p=0.009] and older [OR 1.405, 95% CI 1.006-1.022; p<0.001] participants were more likely to think EMS staff would experience physical violence while at work, table 26.

**Table 25:**
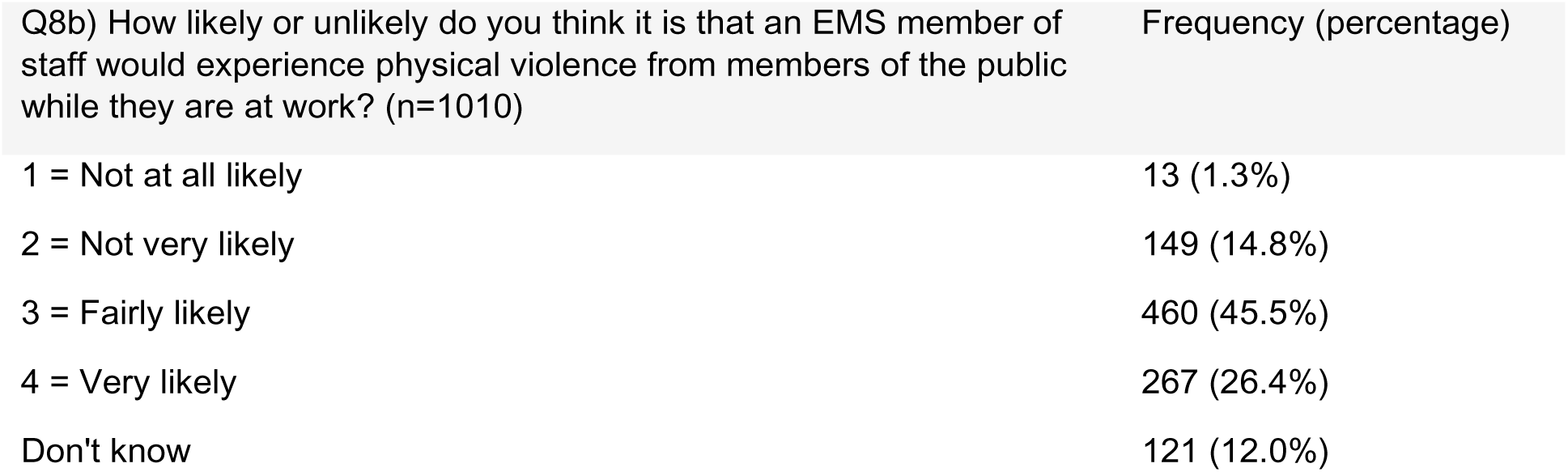
Tabulation of “ How likely or unlikely do you think it is that an EMS member of staff would experience physical violence from members of the public while they are at work?”

**Table 26:**
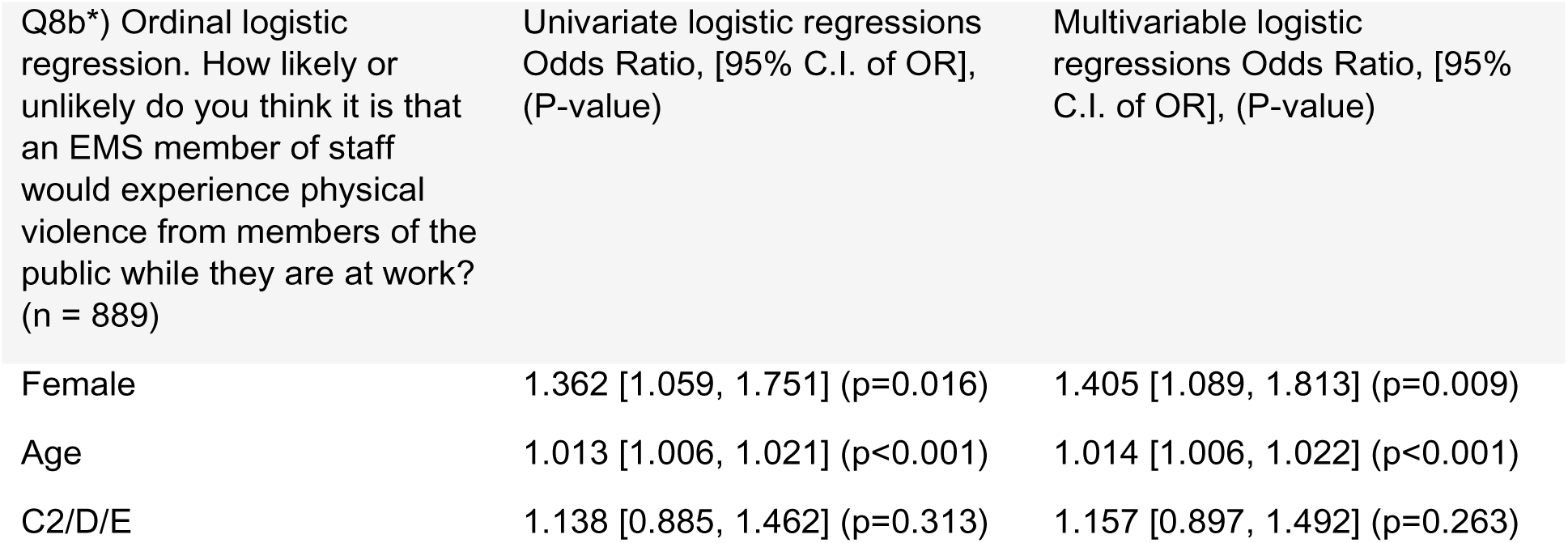
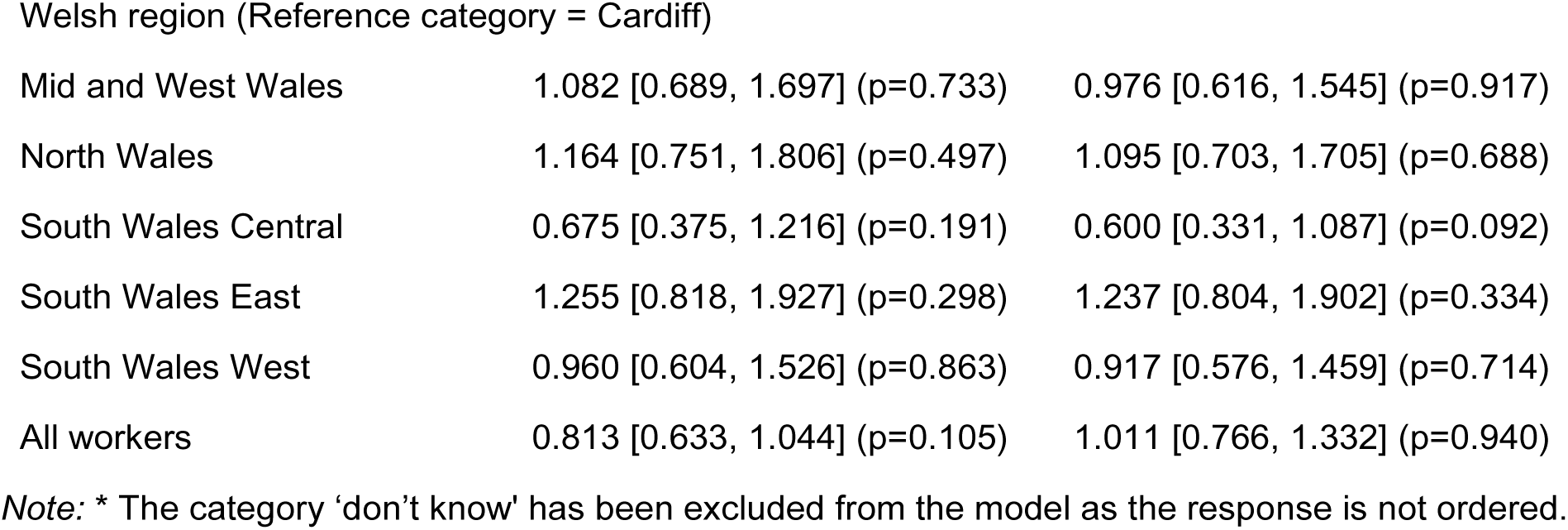
Ordinal logistic regression of “How likely or unlikely do you think it is that an EMS member of staff would experience physical violence from members of the public while they are at work?”

| Q8b*) Ordinal logistic regression. How likely or unlikely do you think it is that an EMS member of staff would experience physical violence from members of the public while they are at work? (n = 889) | Univariate logistic regressions Odds Ratio, [95% C.I. of OR], (P-value) | Multivariable logistic regressions Odds Ratio, [95% C.I. of OR], (P-value) |
| --- | --- | --- |
| Female | 1.362 [1.059, 1.751] (p=0.016) | 1.405 [1.089, 1.813] (p=0.009) |
| Age | 1.013 [1.006, 1.021] (p<0.001) | 1.014 [1.006, 1.022] (p<0.001) |
| C2/D/E | 1.138 [0.885, 1.462] (p=0.313) | 1.157 [0.897, 1.492] (p=0.263) |
| Welsh region (Reference category = Cardiff) |  |  |
| Mid and West Wales | 1.082 [0.689, 1.697] (p=0.733) | 0.976 [0.616, 1.545] (p=0.917) |
| North Wales | 1.164 [0.751, 1.806] (p=0.497) | 1.095 [0.703, 1.705] (p=0.688) |
| South Wales Central | 0.675 [0.375, 1.216] (p=0.191) | 0.600 [0.331, 1.087] (p=0.092) |
| South Wales East | 1.255 [0.818, 1.927] (p=0.298) | 1.237 [0.804, 1.902] (p=0.334) |
| South Wales West | 0.960 [0.604, 1.526] (p=0.863) | 0.917 [0.576, 1.459] (p=0.714) |
| All workers | 0.813 [0.633, 1.044] (p=0.105) | 1.011 [0.766, 1.332] (p=0.940) |

Note: \* The category 'don't know' has been excluded from the model as the response is not ordered.

58.7% of participants thought it to be very, or fairly likely that an EMS member of staff would experience sexual assault/harassment (e.g. use of explicit or implicit sexual overtones, unwanted touching, etc.) from members of the public while they are at work; 25.1% thought it was not at all/very likely with 16.1% responding they did not know, table 27. Women [OR 1.600, 95% CI 1.231,2.080; p<0.001] were more likely to think EMS staff would experience sexual assault/harassment while at work, table 28.

**Table 27:** Tabulation of “How likely or unlikely do you think it is that an EMS member of staff would experience sexual assault/harassment from members of the public while they are at work?”

| Q8c) How likely or unlikely do you think it is that an EMS member of staff would experience sexual assault/harassment from members of the public while they are at work? (n=1010) | Frequency (percentage) |
| --- | --- |
| 1 = Not at all likely | 35 (3.5%) |
| 2 = Not very likely | 219 (21.7%) |
| 3 = Fairly likely | 452 (44.8%) |
| 4 = Very likely | 141 (14.0%) |
| Don't know | 163 (16.1%) |

**Table 28:**
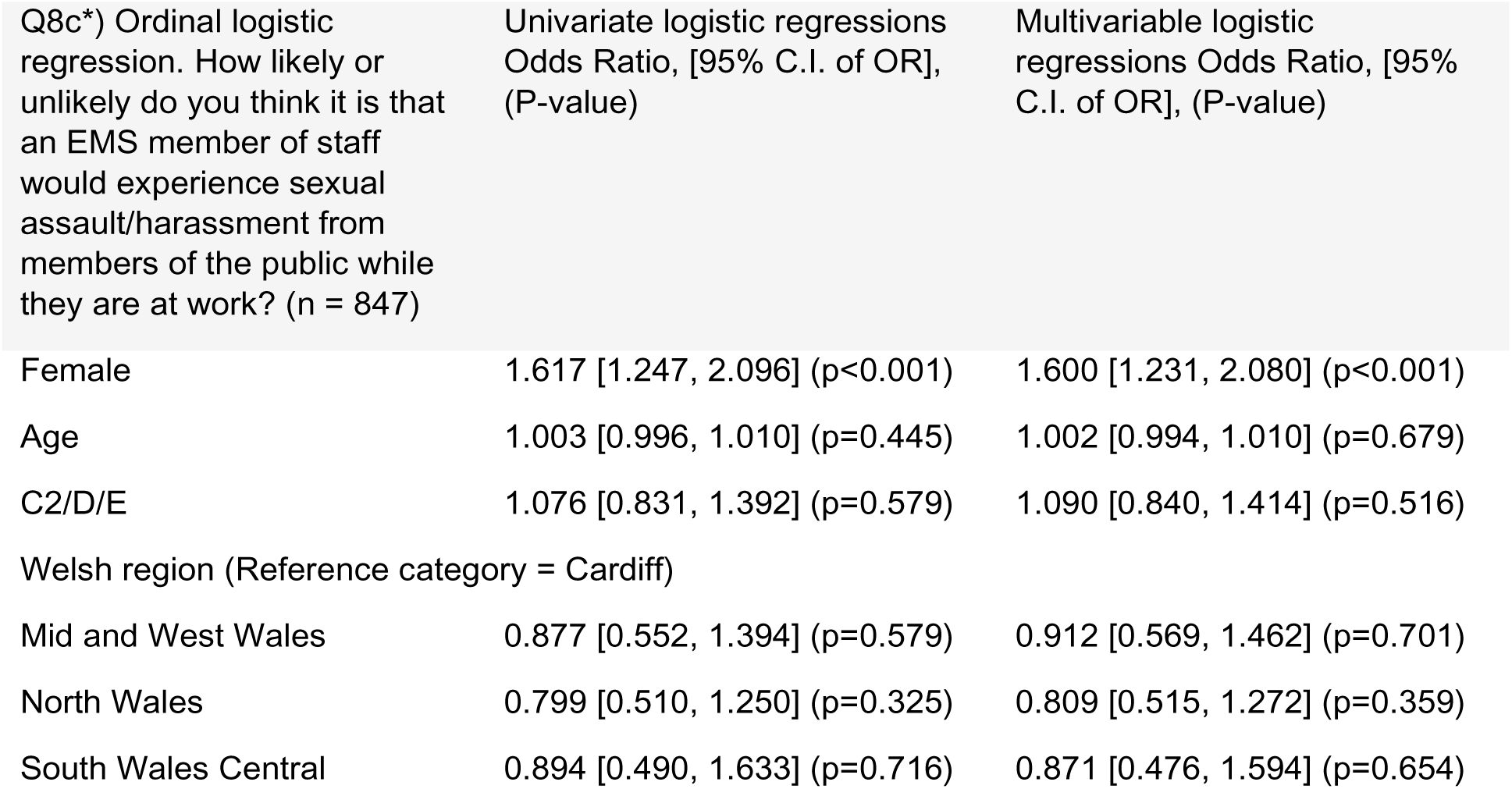
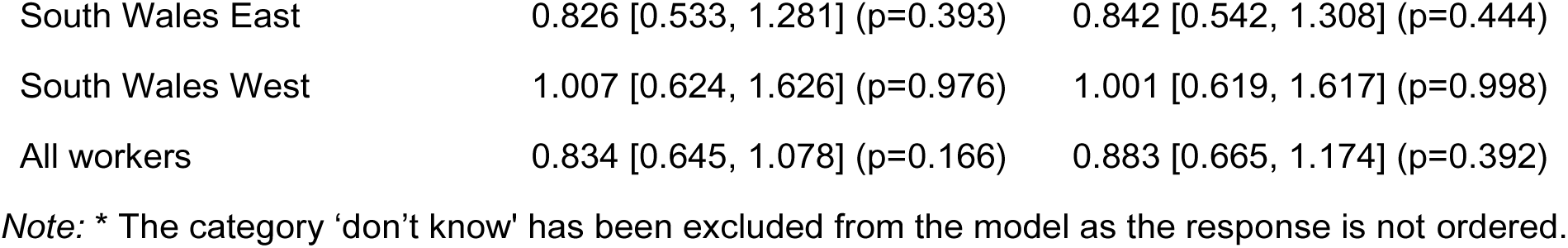
Ordinal logistic regression of “How likely or unlikely do you think it is that an EMS member of staff would experience sexual assault/harassment from members of the public while they are at work?”

### Attitudes to those most likely to direct V&A towards EMS staff

We asked participants how likely members of the public, with varying relations to the patient and EMS staff member, were to be the perpetrator of violent and/or aggressive incidents towards EMS staff. 64.2% of participants thought it was likely that the patient being treated would be violent and/or aggressive, table 29. No characteristics were indicative of a respondent being more likely to think a patient being treated would be violent and/or aggressive towards the EMS staff, table 30. 61.8% of participants thought it was likely that a relative/friend of the patient being treated would be violent and/or aggressive, table 29. Social grades of C2/D/E [OR 1.344, 95% CI 1.033-1.749, p=0.028] were more likely to think a relative/friend of the patient being treated would be violent and/pr aggressive towards the EMS staff, table 31. 31.0% of participants thought it was likely that a bystander/stranger with no relation to the patient or EMS staff member would be violent and/or aggressive, table 29. No characteristics were indicative of a respondent being more likely to think a bystander/stranger, with no relation to the patient or EMS staff, would be violent and/or aggressive towards the EMS staff, table 32.

**Table 29:** Tabulation of “How likely or unlikely do you think it is that an EMS member of staff would experience the following forms of violence and/or aggression from members of the public while they are at work?”

| Q9) How likely or unlikely do you think the following people are to be violent and/or aggressive towards EMS staff while they are trying to treat a patient? (n=1010) | Frequency (percentage) |  |  |
| --- | --- | --- | --- |
|  | Q9a) The patient being treated by the EMS staff | Q9b) A relative/friend of the patient being treated by EMS staff | Q9c) A bystander/stranger with no relation to the patient or EMS staff |
| 1 = Not at all likely | 32 (3.2%) | 35 (3.5%) | 99 (9.8%) |
| 2 = Not very likely | 200 (19.8%) | 202 (20.0%) | 434 (43.0%) |
| 3 = Fairly likely | 455 (45.0%) | 489 (48.4%) | 250 (24.8%) |
| 4 = Very likely | 193 (19.1%) | 135 (13.4%) | 63 (6.2%) |
| Don't know | 130 (12.9%) | 149 (14.8%) | 164 (16.2%) |

**Table 30:** Ordinal logistic regression of “How likely or unlikely do you think the patient being treated by the EMS staff are to be violent and/or aggressive towards EMS staff while they are trying to treat a patient?”

| Q9a*) Ordered logistic regression. How likely or unlikely do you think **the patient being treated by the EMS staff** are to be violent and/or aggressive towards EMS staff while they are trying to treat a patient? (n=880) | Univariate logistic regressions Odds Ratio, [95% C.I. of OR], (P-value) | Multivariable logistic regressions Odds Ratio, [95% C.I. of OR], (P-value) |
| --- | --- | --- |
| Female | 1.214 [0.945, 1.560] (0.129) | 1.204 [0.935, 1.551] (0.150) |
| Age | 0.998 [0.991, 1.005] (0.518) | 0.998 [0.990, 1.005] (0.532) |
| C2/D/E | 1.227 [0.955, 1.576] (0.109) | 1.241 [0.964, 1.598] (0.094) |
| Welsh region (Reference category = Cardiff) |  |  |
| Mid and West Wales | 0.843 [0.540, 1.314] (p=0.450) | 0.852 [0.543, 1.336] (p=0.485) |
| North Wales | 1.196 [0.776, 1.844] (p=0.417) | 1.187 [0.769, 1.833] (p=0.439) |
| South Wales Central | 0.960 [0.533, 1.730] (p=0.892) | 0.918 [0.508, 1.659] (p=0.778) |
| South Wales East | 1.114 [0.729, 1.701] (p=0.618) | 1.103 [0.722, 1.685] (p=0.649) |
| South Wales West | 1.038 [0.655, 1.645] (p=0.874) | 1.019 [0.643, 1.616] (p=0.936) |
| ALL WORKERS (NET) | 0.958 [0.746, 1.230] (p=0.738) | 0.940 [0.714, 1.237] (p=0.660) |

Note: \* The category 'don't know' has been excluded from the model as the response is not ordered.

**Table 31:** Ordinal logistic regression of “How likely or unlikely do you think a relative/friend of the patient being treated by EMS staff are to be violent and/or aggressive towards EMS staff while they are trying to treat a patient?”

| Q9b*) Ordered logistic regression. How likely or unlikely do you think **a relative/ friend of the patient being treated by EMS staff** are to be violent and/or aggressive towards EMS staff while they are trying to treat a patient? (n=861) | Univariate logistic regressions Odds Ratio, [95% C.I. of OR], (P-value) | Multivariable logistic regressions Odds Ratio, [95% C.I. of OR], (P-value) |
| --- | --- | --- |
| Female | 1.125 [0.867, 1.459] (p=0.375) | 1.169 [0.899, 1.521] (p=0.244) |
| Age | 1.003 [0.995, 1.010] (p=0.491) | 1.004 [0.995, 1.012] (p=0.387) |
| C2/D/E | 1.329 [1.024, 1.724] (p=0.032) | 1.344 [1.033, 1.749] (p=0.028) |
| Welsh region (Reference category = Cardiff) |  |  |
| Mid and West Wales | 1.193 [0.748, 1.902] (p=0.459) | 1.169 [0.727, 1.880] (p=0.520) |
| North Wales | 1.192 [0.759, 1.870] (p=0.445) | 1.171 [0.745, 1.841] (p=0.493) |
| South Wales Central | 1.100 [0.596, 2.029] (p=0.761) | 1.055 [0.571, 1.951] (p=0.864) |
| South Wales East | 0.921 [0.593, 1.432] (p=0.715) | 0.903 [0.581, 1.405] (p=0.652) |
| South Wales West | 1.209 [0.750, 1.949] (p=0.436) | 1.178 [0.730, 1.901] (p=0.502) |
| ALL WORKERS (NET) | 1.044 [0.806, 1.352] (p=0.743) | 1.172 [0.880, 1.562] (p=0.277) |

Note: \* The category 'don't know' has been excluded from the model as the response is not ordered.

**Table 32:**
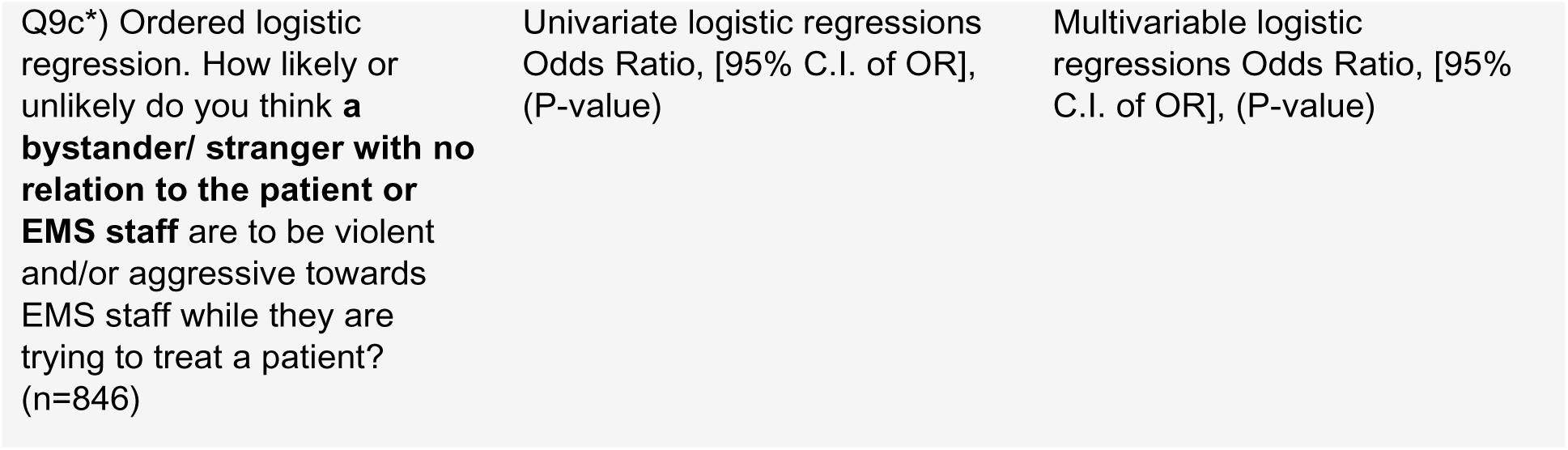
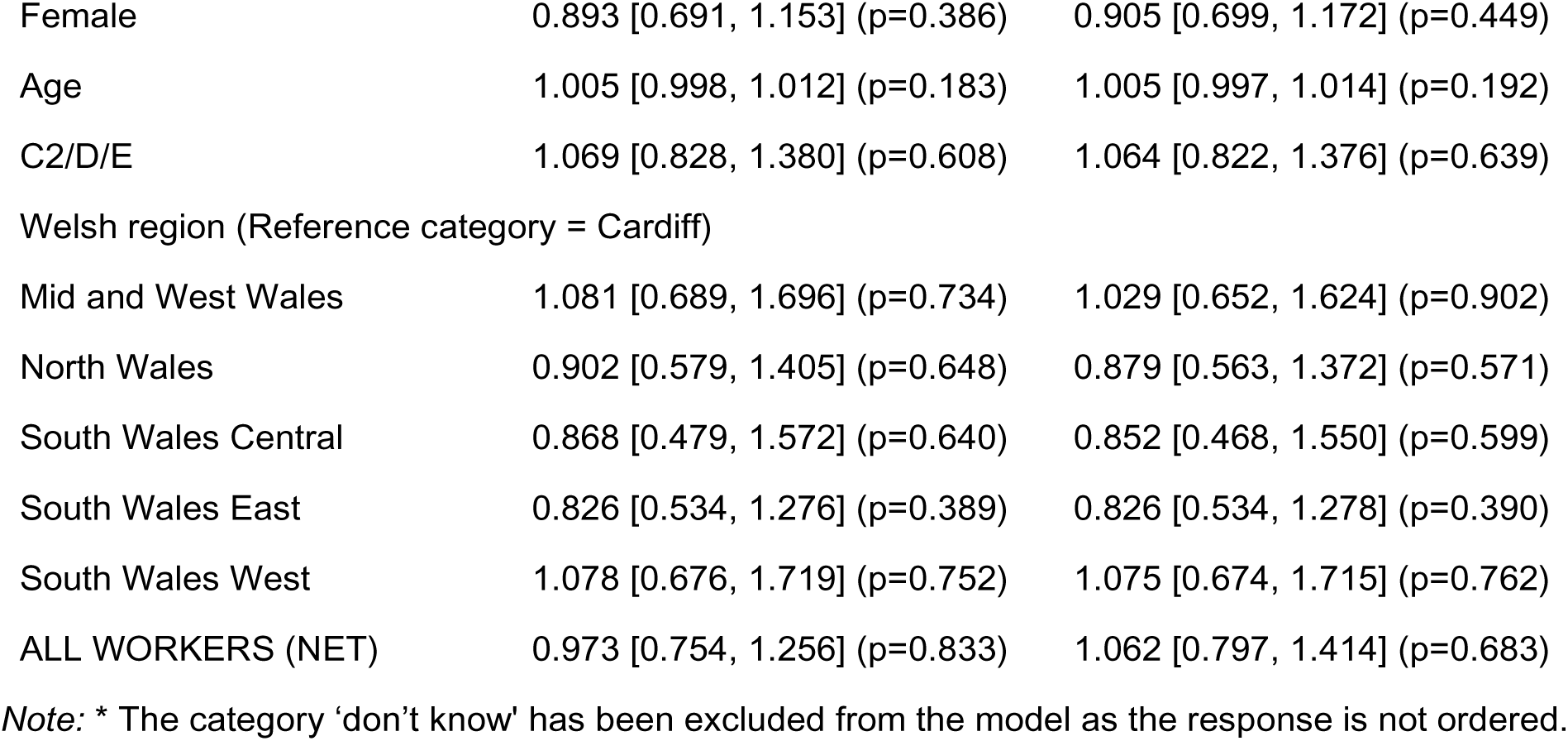
Ordinal logistic regression of “How likely or unlikely do you think a bystander/stranger with no relation to the patient or EMS staff are to be violent and/or aggressive towards EMS staff while they are trying to treat a patient? “.

| Q9c*) Ordered logistic regression. How likely or unlikely do you think **a bystander/ stranger with no relation to the patient or EMS staff** are to be violent and/or aggressive towards EMS staff while they are trying to treat a patient? (n=846) | Univariate logistic regressions Odds Ratio, [95% C.I. of OR], (P-value) | Multivariable logistic regressions Odds Ratio, [95% C.I. of OR], (P-value) |
| --- | --- | --- |
| Female | 0.893 [0.691, 1.153] (p=0.386) | 0.905 [0.699, 1.172] (p=0.449) |
| Age | 1.005 [0.998, 1.012] (p=0.183) | 1.005 [0.997, 1.014] (p=0.192) |
| C2/D/E | 1.069 [0.828, 1.380] (p=0.608) | 1.064 [0.822, 1.376] (p=0.639) |
| Welsh region (Reference category = Cardiff) |  |  |
| Mid and West Wales | 1.081 [0.689, 1.696] (p=0.734) | 1.029 [0.652, 1.624] (p=0.902) |
| North Wales | 0.902 [0.579, 1.405] (p=0.648) | 0.879 [0.563, 1.372] (p=0.571) |
| South Wales Central | 0.868 [0.479, 1.572] (p=0.640) | 0.852 [0.468, 1.550] (p=0.599) |
| South Wales East | 0.826 [0.534, 1.276] (p=0.389) | 0.826 [0.534, 1.278] (p=0.390) |
| South Wales West | 1.078 [0.676, 1.719] (p=0.752) | 1.075 [0.674, 1.715] (p=0.762) |
| ALL WORKERS (NET) | 0.973 [0.754, 1.256] (p=0.833) | 1.062 [0.797, 1.414] (p=0.683) |

Note: \* The category 'don't know' has been excluded from the model as the response is not ordered.

### Attitudes to factors of V&A directed towards EMS staff

We explored participants’ perceptions of how likely a factor was to contribute to violent and/or aggressive incidents towards EMS staff. 92.4% thought intoxication with alcohol was likely to contribute to violent and/or aggressive incidents, table 33. Participants from South Wales Central, compared to Cardiff, [OR 0.481, 95% CI 0.240-0.963; p=0.039] were less likely to think intoxication with alcohol would contribute to violent and/or aggressive incidents, table 34. 90.5% thought intoxication with drugs was likely contribute to violent and/or aggressive incidents, table 33. Females [OR 1.400, 95% CI 1.041-1.883; p=0.026] and older [OR 1.015, 95% CI 1.005-1.024; p=0.002] participants were more likely to think intoxication with drugs would contribute to violent and/or aggressive incidents, table 35. 84.3% thought an altered mental status following illness and/or injury was likely contributed to violent and/or aggressive incidents, table 33. No characteristics were indicative of a respondent being more likely to think an altered mental status following illness and/or injury was likely to contributed to violent and/or aggressive incidents, table 36.

**Table 33:**
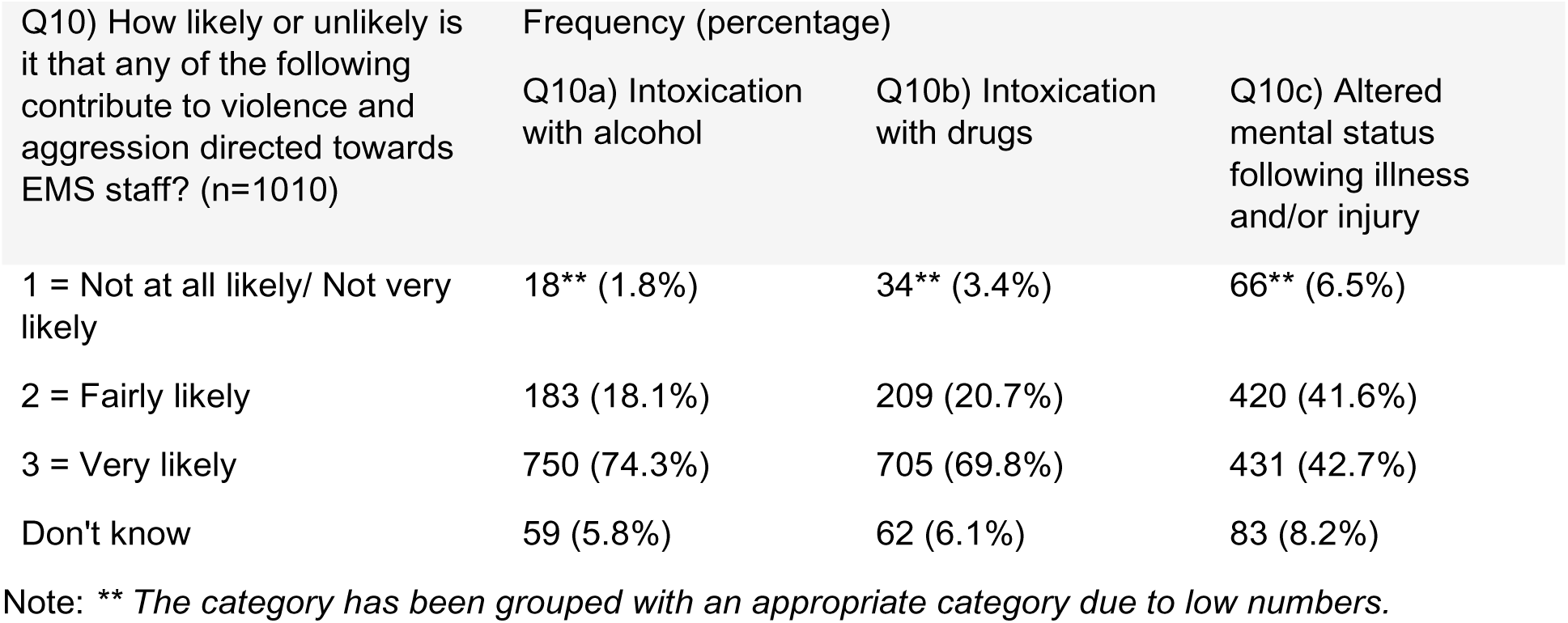
Tabulation of “How likely or unlikely is it that any of the following contribute to violence and aggression directed towards EMS staff?”

| Q10) How likely or unlikely is it that any of the following contribute to violence and aggression directed towards EMS staff? (n=1010) | Frequency (percentage) |  |  |
| --- | --- | --- | --- |
|  | Q10a) Intoxication with alcohol | Q10b) Intoxication with drugs | Q10c) Altered mental status following illness and/or injury |
| 1 = Not at all likely/ Not very likely | 18** (1.8%) | 34** (3.4%) | 66** (6.5%) |
| 2 = Fairly likely | 183 (18.1%) | 209 (20.7%) | 420 (41.6%) |
| 3 = Very likely | 750 (74.3%) | 705 (69.8%) | 431 (42.7%) |
| Don't know | 59 (5.8%) | 62 (6.1%) | 83 (8.2%) |

Note: \*\* The category has been grouped with an appropriate category due to low numbers.

**Table 34:**
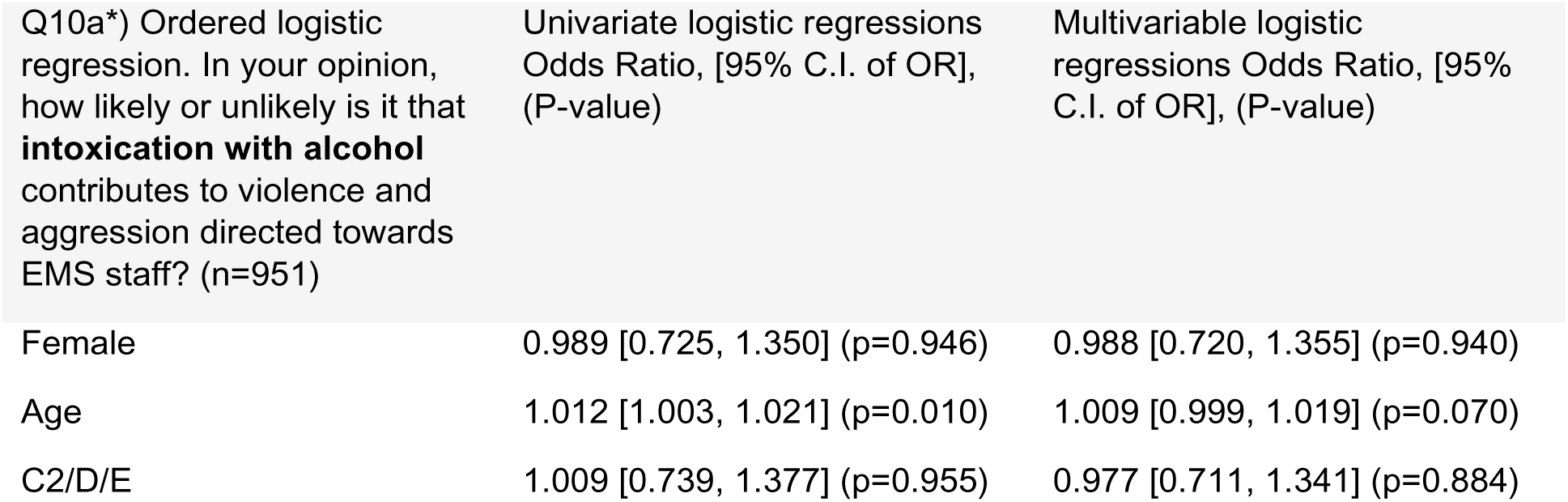
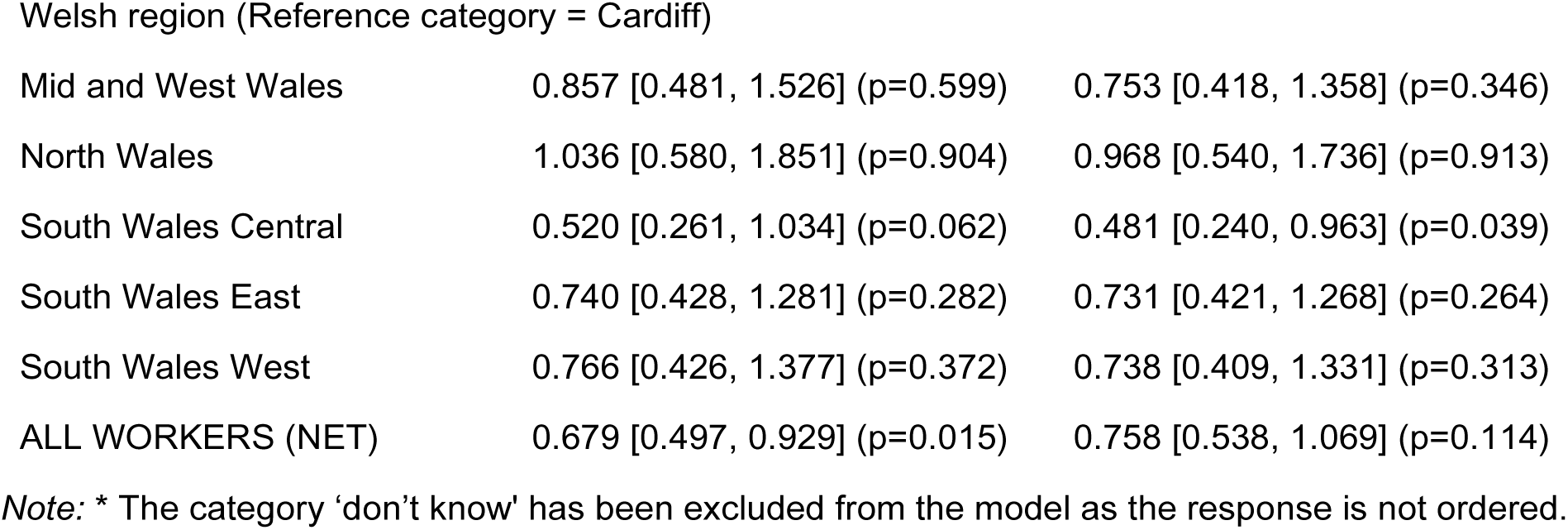
Ordinal logistic regression of “In your opinion, how likely or unlikely is it that intoxication with alcohol contributes to violence and aggression directed towards EMS staff?”

| Q10a*) Ordered logistic regression. In your opinion, how likely or unlikely is it that **intoxication with alcohol** contributes to violence and aggression directed towards EMS staff? (n=951) | Univariate logistic regressions Odds Ratio, [95% C.I. of OR], (P-value) | Multivariable logistic regressions Odds Ratio, [95% C.I. of OR], (P-value) |
| --- | --- | --- |
| Female | 0.989 [0.725, 1.350] (p=0.946) | 0.988 [0.720, 1.355] (p=0.940) |
| Age | 1.012 [1.003, 1.021] (p=0.010) | 1.009 [0.999, 1.019] (p=0.070) |
| C2/D/E | 1.009 [0.739, 1.377] (p=0.955) | 0.977 [0.711, 1.341] (p=0.884) |
| Welsh region (Reference category = Cardiff) |  |  |
| Mid and West Wales | 0.857 [0.481, 1.526] (p=0.599) | 0.753 [0.418, 1.358] (p=0.346) |
| North Wales | 1.036 [0.580, 1.851] (p=0.904) | 0.968 [0.540, 1.736] (p=0.913) |
| South Wales Central | 0.520 [0.261, 1.034] (p=0.062) | 0.481 [0.240, 0.963] (p=0.039) |
| South Wales East | 0.740 [0.428, 1.281] (p=0.282) | 0.731 [0.421, 1.268] (p=0.264) |
| South Wales West | 0.766 [0.426, 1.377] (p=0.372) | 0.738 [0.409, 1.331] (p=0.313) |
| ALL WORKERS (NET) | 0.679 [0.497, 0.929] (p=0.015) | 0.758 [0.538, 1.069] (p=0.114) |

Note: \* The category 'don't know' has been excluded from the model as the response is not ordered.

**Table 35:**
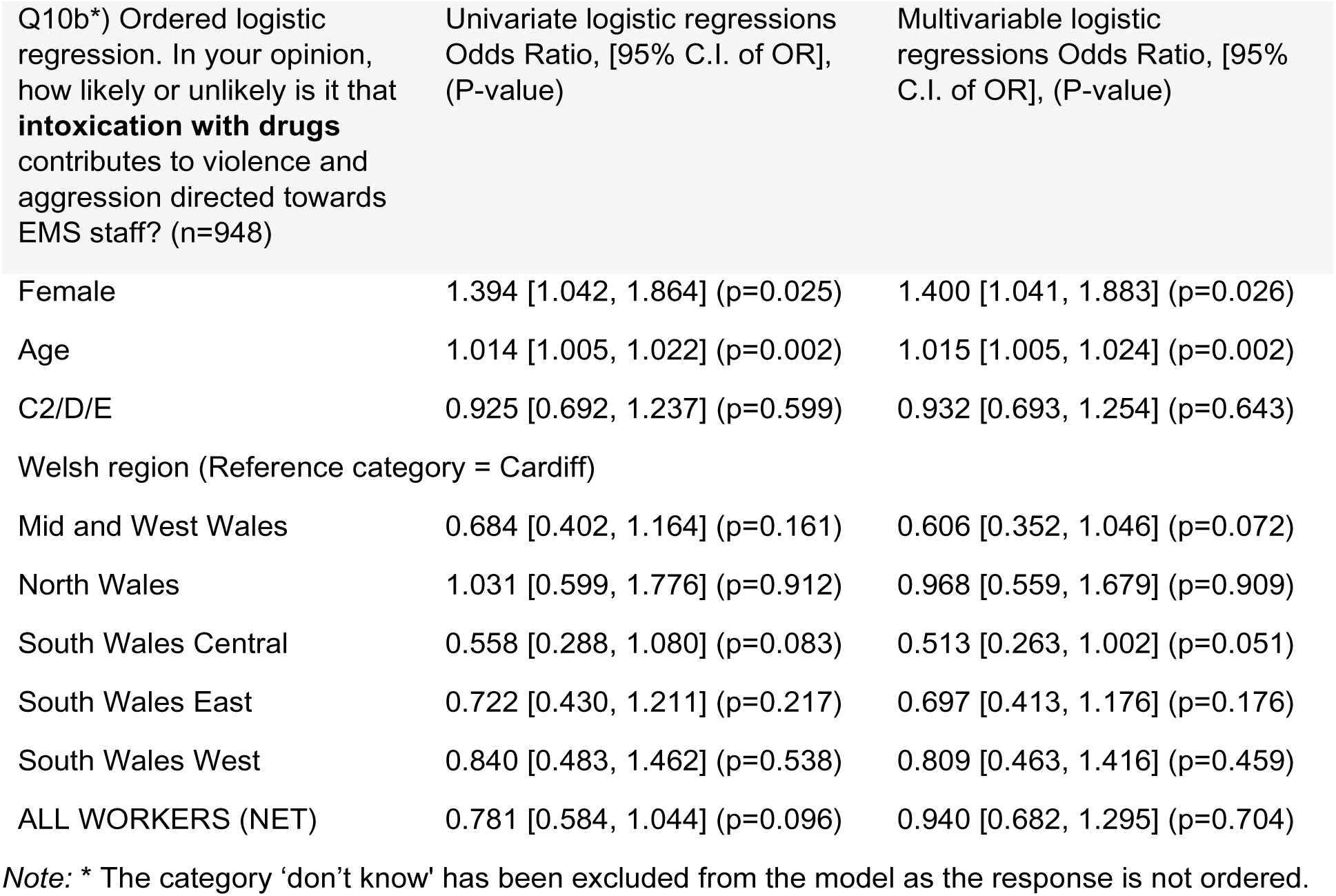
Ordinal logistic regression of “In your opinion, how likely or unlikely is it that intoxication with drugs contributes to violence and aggression directed towards EMS staff?”

| Q10b*) Ordered logistic regression. In your opinion, how likely or unlikely is it that **intoxication with drugs** contributes to violence and aggression directed towards EMS staff? (n=948) | Univariate logistic regressions Odds Ratio, [95% C.I. of OR], (P-value) | Multivariable logistic regressions Odds Ratio, [95% C.I. of OR], (P-value) |
| --- | --- | --- |
| Female | 1.394 [1.042, 1.864] (p=0.025) | 1.400 [1.041, 1.883] (p=0.026) |
| Age | 1.014 [1.005, 1.022] (p=0.002) | 1.015 [1.005, 1.024] (p=0.002) |
| C2/D/E | 0.925 [0.692, 1.237] (p=0.599) | 0.932 [0.693, 1.254] (p=0.643) |
| Welsh region (Reference category = Cardiff) |  |  |
| Mid and West Wales | 0.684 [0.402, 1.164] (p=0.161) | 0.606 [0.352, 1.046] (p=0.072) |
| North Wales | 1.031 [0.599, 1.776] (p=0.912) | 0.968 [0.559, 1.679] (p=0.909) |
| South Wales Central | 0.558 [0.288, 1.080] (p=0.083) | 0.513 [0.263, 1.002] (p=0.051) |
| South Wales East | 0.722 [0.430, 1.211] (p=0.217) | 0.697 [0.413, 1.176] (p=0.176) |
| South Wales West | 0.840 [0.483, 1.462] (p=0.538) | 0.809 [0.463, 1.416] (p=0.459) |
| ALL WORKERS (NET) | 0.781 [0.584, 1.044] (p=0.096) | 0.940 [0.682, 1.295] (p=0.704) |

Note: \* The category 'don't know' has been excluded from the model as the response is not ordered.

**Table 36:**
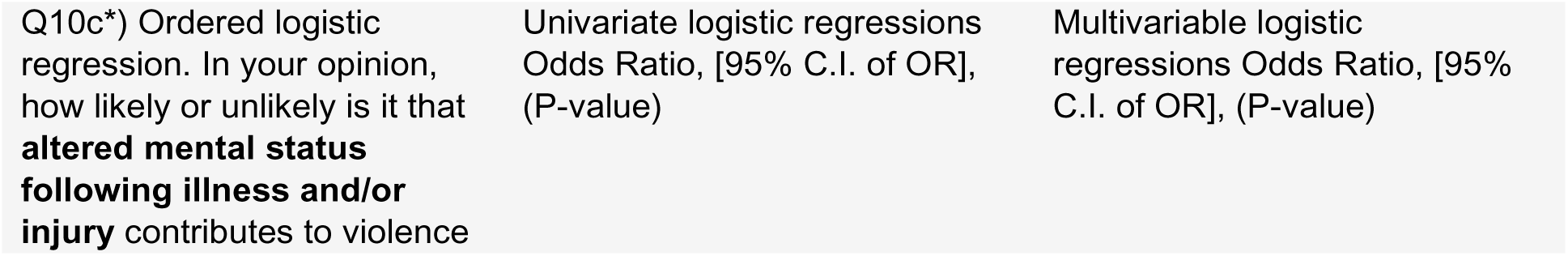
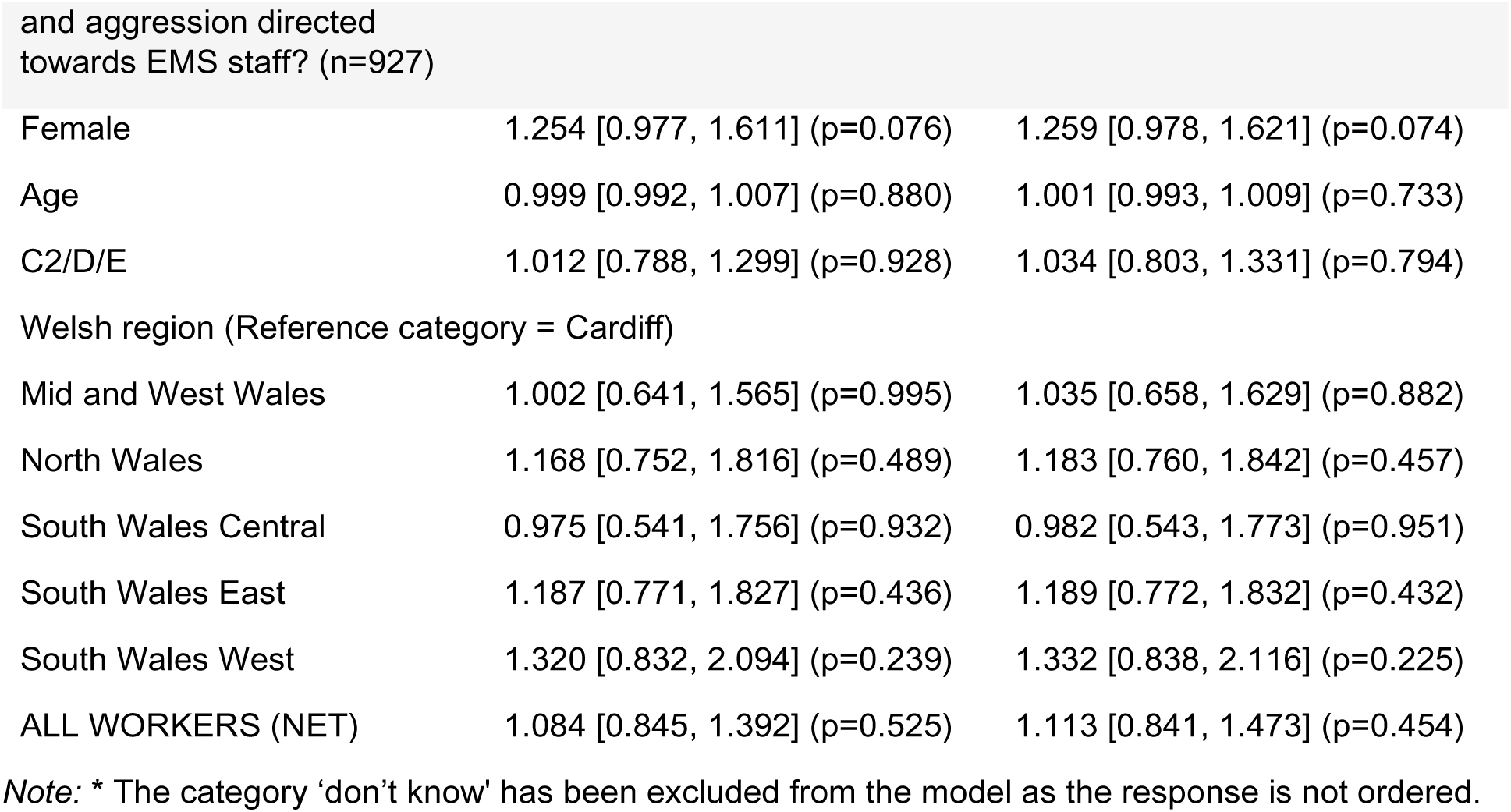
Ordinal logistic regression of “In your opinion, how likely or unlikely is it that altered mental status following illness and/or injury contributes to violence and aggression directed towards EMS staff?”

| Q10c*) Ordered logistic regression. In your opinion, how likely or unlikely is it that **altered mental status following illness and/or injury** contributes to violence | Univariate logistic regressions Odds Ratio, [95% C.I. of OR], (P-value) | Multivariable logistic regressions Odds Ratio, [95% C.I. of OR], (P-value) |
| --- | --- | --- |
| Female | 1.254 [0.977, 1.611] (p=0.076) | 1.259 [0.978, 1.621] (p=0.074) |
| Age | 0.999 [0.992, 1.007] (p=0.880) | 1.001 [0.993, 1.009] (p=0.733) |
| C2/D/E | 1.012 [0.788, 1.299] (p=0.928) | 1.034 [0.803, 1.331] (p=0.794) |
| Welsh region (Reference category = Cardiff) |  |  |
| Mid and West Wales | 1.002 [0.641, 1.565] (p=0.995) | 1.035 [0.658, 1.629] (p=0.882) |
| North Wales | 1.168 [0.752, 1.816] (p=0.489) | 1.183 [0.760, 1.842] (p=0.457) |
| South Wales Central | 0.975 [0.541, 1.756] (p=0.932) | 0.982 [0.543, 1.773] (p=0.951) |
| South Wales East | 1.187 [0.771, 1.827] (p=0.436) | 1.189 [0.772, 1.832] (p=0.432) |
| South Wales West | 1.320 [0.832, 2.094] (p=0.239) | 1.332 [0.838, 2.116] (p=0.225) |
| ALL WORKERS (NET) | 1.084 [0.845, 1.392] (p=0.525) | 1.113 [0.841, 1.473] (p=0.454) |

Note: \* The category 'don't know' has been excluded from the model as the response is not ordered.

### Awareness of, and attitudes to legislation and media campaigns related to V&A directed towards EMS staff

We asked participants if, before taking the survey, they were aware of the Assaults on Emergency Workers (Offences) Act of 2018; 22% reported being aware of the act. Table 37; men [OR 0.621, 95% CI 0.459-0.842; p=0.002] were more likely to be aware of the act, table 38.

**Table 37:** Tabulation of “Were you aware of the Assaults on Emergency Workers (Offences) Act of 2018?”

| Q11) Were you aware of the Assaults on Emergency Workers (Offences) Act of 2018? (n=1010) | Frequency (percentage) |
| --- | --- |
| No, I wasn't | 788 (78.0%) |
| Yes, I was | 222 (22.0%) |

**Table 38:**
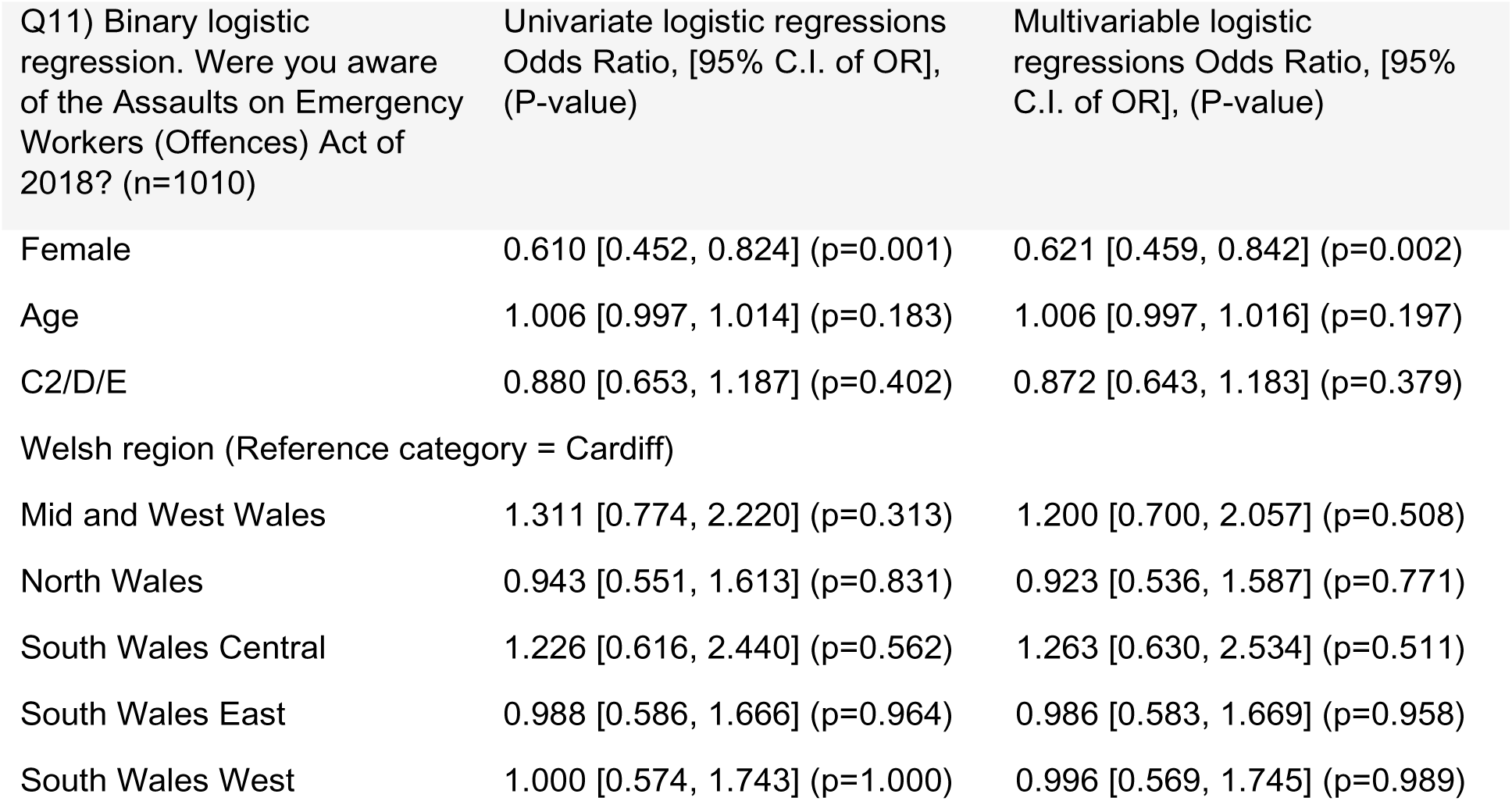
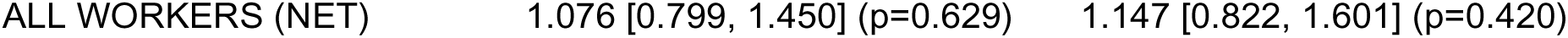
Binary logistic regression of “Were you aware of the Assaults on Emergency Workers (Offences) Act of 2018?”

The Assaults on Emergency Workers (Offences) Act (2018) was described to participants in the questionnaire (appendix 1). We asked how likely participants thought the Assault on Emergency Workers (Offences) Act (2018) would be at deterring members of the public. 75.2% thought it was unlikely that those intoxicated with alcohol would be deterred from being violent and/or aggressive towards EMS staff and 15.4% thought it was likely, table 39. Younger [OR 0.987, 95% CI 0.979-0.994; p=0.001] participants were more likely to think those intoxicated with alcohol would be deterred, table 40. 75.2% thought it was unlikely that those intoxicated with drugs would be deterred from being violent and/or aggressive towards EMS staff and 15.0% thought it was likely, table 39. Younger [OR 0.992, 95% CI 0.984-0.999; p=0.033] and participants with a social grade of C2/D/E [OR 1.315, 95% CI 1.024-1.689; p=0.032] were more likely to think those intoxicated with drugs would be deterred, table 41. 75.6% thought it was unlikely that those with an altered mental status following illness and/or injury would be deterred from being violent and/or aggressive towards EMS staff and 13.0% it was like, table 39. Younger [OR 0.989, 95% CI 0.981-0.997; p=0.006] participants were more likely to think those with an altered mental status following illness and/or injury would be deterred, table 42. 42.2% thought it was unlikely that other members of the public would be deterred from being violent and/or aggressive towards EMS staff and 42.9% thought it was likely, table 39. Younger [OR 0.991, 95% CI 0.983-0.999; p=0.029] participants were more likely to think other members of the public would be deterred, table 43.

**Table 39:**
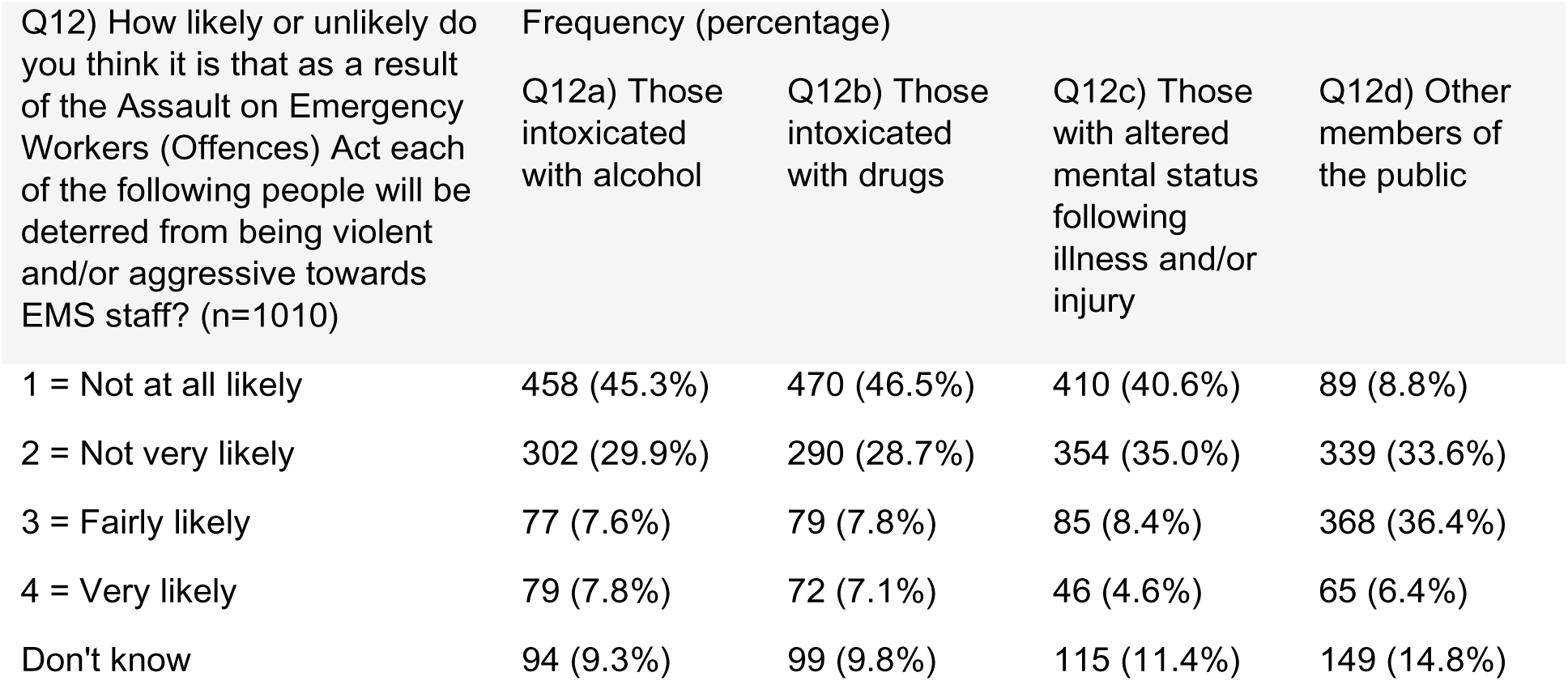
Tabulation of “How likely or unlikely do you think it is that as a result of the Assault on Emergency Workers (Offences) Act each of the following people will be deterred from being violent and/or aggressive towards EMS staff?”

| Q12) How likely or unlikely do you think it is that as a result of the Assault on Emergency Workers (Offences) Act each of the following people will be deterred from being violent and/or aggressive towards EMS staff? (n=1010) | Frequency (percentage) |  |  |  |
| --- | --- | --- | --- | --- |
|  | Q12a) Those intoxicated with alcohol | Q12b) Those intoxicated with drugs | Q12c) Those with altered mental status following illness and/or injury | Q12d) Other members of the public |
| 1 = Not at all likely | 458 (45.3%) | 470 (46.5%) | 410 (40.6%) | 89 (8.8%) |
| 2 = Not very likely | 302 (29.9%) | 290 (28.7%) | 354 (35.0%) | 339 (33.6%) |
| 3 = Fairly likely | 77 (7.6%) | 79 (7.8%) | 85 (8.4%) | 368 (36.4%) |
| 4 = Very likely | 79 (7.8%) | 72 (7.1%) | 46 (4.6%) | 65 (6.4%) |
| Don't know | 94 (9.3%) | 99 (9.8%) | 115 (11.4%) | 149 (14.8%) |

**Table 40:**
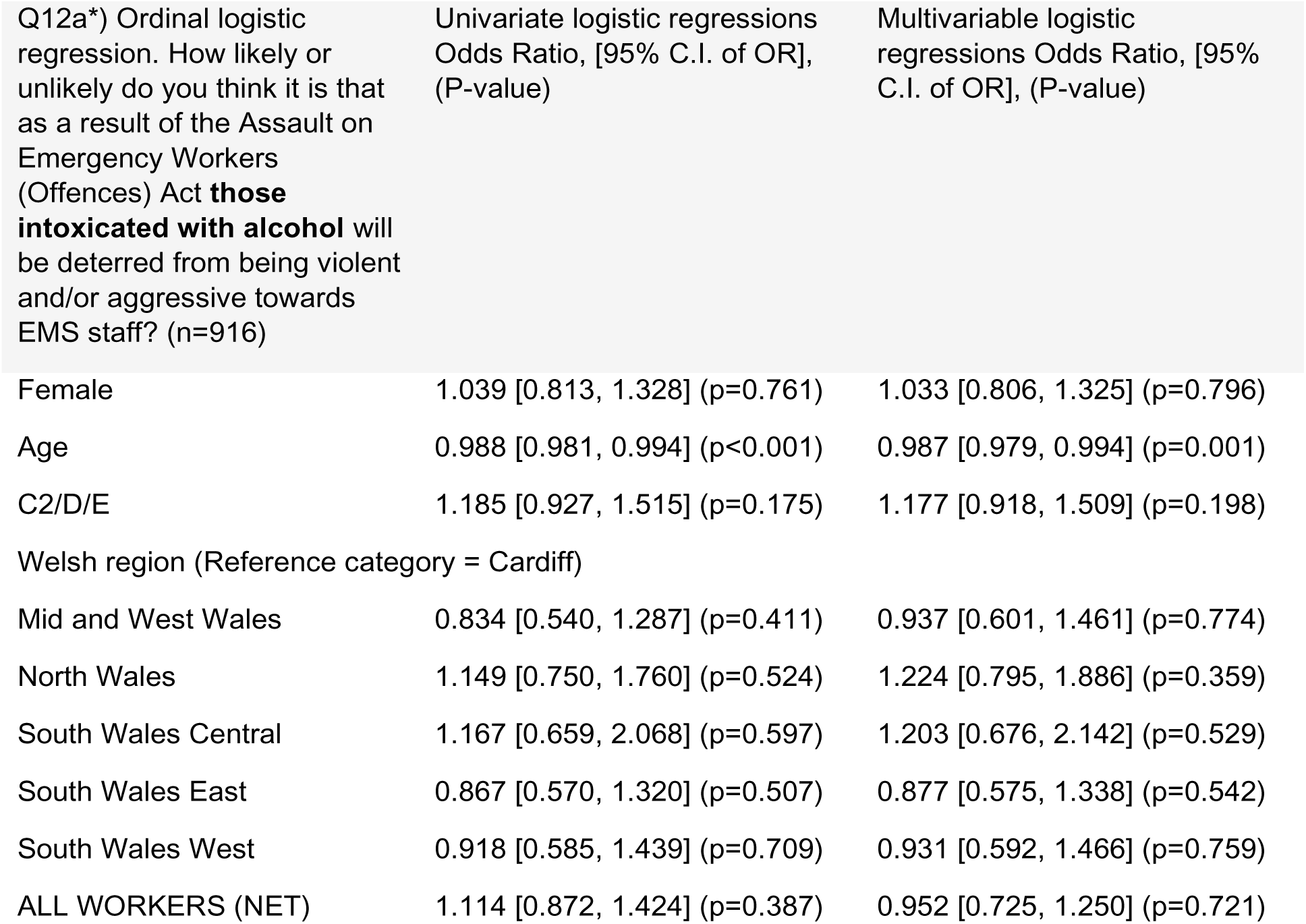
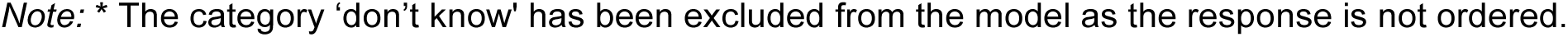
Ordinal logistic regression of “How likely or unlikely do you think it is that as a result of the Assault on Emergency Workers (Offences) Act those intoxicated with alcohol will be deterred from being violent and/or aggressive towards EMS staff?”

**Table 41:**
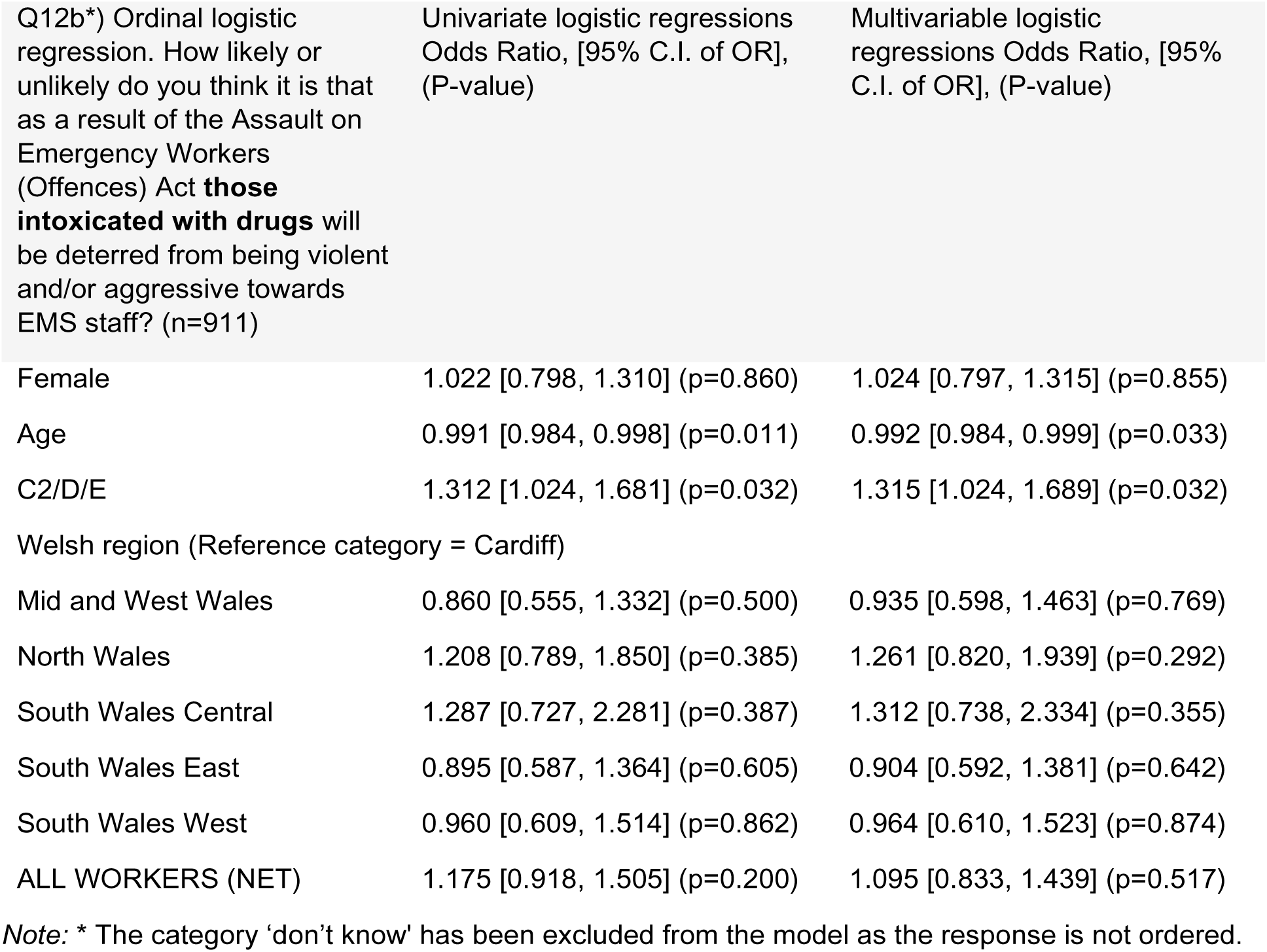
Ordinal logistic regression of “How likely or unlikely do you think it is that as a result of the Assault on Emergency Workers (Offences) Act those intoxicated with drugs will be deterred from being violent and/or aggressive towards EMS staff?”

**Table 42:** Ordinal logistic regression of “How likely or unlikely do you think it is that as a result of the Assault on Emergency Workers (Offences) Act those with altered mental status following illness and/or injury will be deterred from being violent and/or aggressive towards EMS staff?”

| Q12c*) Ordinal logistic regression. How likely or unlikely do you think it is that as a result of the Assault on Emergency Workers (Offences) Act **those with altered mental status following illness and/or injury** will be deterred from being violent and/or aggressive towards EMS staff? (n=895) | Univariate logistic regressions Odds Ratio, [95% C.I. of OR], (P-value) | Multivariable logistic regressions Odds Ratio, [95% C.I. of OR], (P-value) |
| --- | --- | --- |
| Female | 1.096 [0.855, 1.404] (p=0.470) | 1.089 [0.847, 1.400] (p=0.505) |
| Age | 0.988 [0.981, 0.995] (p=0.001) | 0.989 [0.981, 0.997] (p=0.006) |
| C2/D/E | 1.103 [0.861, 1.413] (p=0.440) | 1.116 [0.869, 1.434] (p=0.390) |
| Welsh region (Reference category = Cardiff) |  |  |
| Mid and West Wales | 0.782 [0.504, 1.211] (p=0.270) | 0.885 [0.566, 1.383] (p=0.591) |
| North Wales | 1.172 [0.764, 1.800] (p=0.467) | 1.252 [0.813, 1.929] (p=0.308) |
| South Wales Central | 1.253 [0.695, 2.257] (p=0.454) | 1.322 [0.732, 2.389] (p=0.355) |
| South Wales East | 0.903 [0.595, 1.373] (p=0.634) | 0.915 [0.601, 1.392] (p=0.677) |
| South Wales West | 1.131 [0.721, 1.774] (p=0.593) | 1.157 [0.736, 1.821] (p=0.528) |
| ALL WORKERS (NET) | 1.268 [0.990, 1.626] (p=0.060) | 1.108 [0.842, 1.459] (p=0.465) |

Note: \* The category 'don't know' has been excluded from the model as the response is not ordered.

**Table 43:** Ordinal logistic regression of “How likely or unlikely do you think it is that as a result of the Assault on Emergency Workers (Offences) Act other members of the public will be deterred from being violent and/or aggressive towards EMS staff?”

| Q12d*) Ordinal logistic regression. How likely or unlikely do you think it is that as a result of the Assault on Emergency Workers (Offences) Act **other members of the public** will be deterred from being violent and/or aggressive towards EMS staff? (n=861) | Univariate logistic regressions Odds Ratio, [95% C.I. of OR], (P-value) | Multivariable logistic regressions Odds Ratio, [95% C.I. of OR], (P-value) |
| --- | --- | --- |
| Female | 0.913 [0.711, 1.173] (p=0.477) | 0.904 [0.702, 1.165] (p=0.436) |
| Age | 0.994 [0.987, 1.001] (p=0.080) | 0.991 [0.983, 0.999] (p=0.029) |
| C2/D/E | 0.866 [0.674, 1.113] (p=0.261) | 0.850 [0.660, 1.094] (p=0.208) |
| Welsh region (Reference category = Cardiff) |  |  |
| Mid and West Wales | 0.933 [0.602, 1.447] (p=0.758) | 1.007 [0.645, 1.575] (p=0.974) |
| North Wales | 0.852 [0.553, 1.313] (p=0.468) | 0.886 [0.573, 1.369] (p=0.585) |
| South Wales Central | 0.842 [0.465, 1.524] (p=0.570) | 0.869 [0.479, 1.576] (p=0.645) |
| South Wales East | 0.795 [0.518, 1.220] (p=0.294) | 0.804 [0.524, 1.234] (p=0.319) |
| South Wales West | 0.807 [0.512, 1.272] (p=0.356) | 0.829 [0.526, 1.308] (p=0.421) |
| ALL WORKERS (NET) | 0.991 [0.772, 1.273] (p=0.946) | 0.857 [0.648, 1.133] (p=0.278) |

Note: \* The category 'don't know' has been excluded from the model as the response is not ordered.

66.5% agreed with the statement, “as a result of the Act, those who are violent and/or assault EMS staff will receive harsher sentences” and 7.5% disagreed with the statement, table 44. Older [OR 1.018, 95% CI 1.010-1.026; p<0.001] participants agreed more with the statement that those who are violent and/or assault EMS staff will receive harder sentences.

**Table 44:**
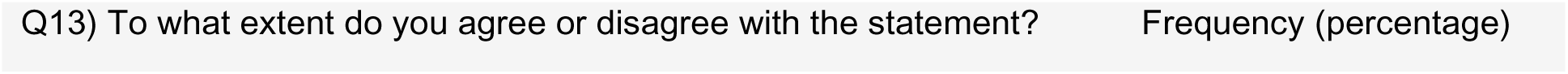
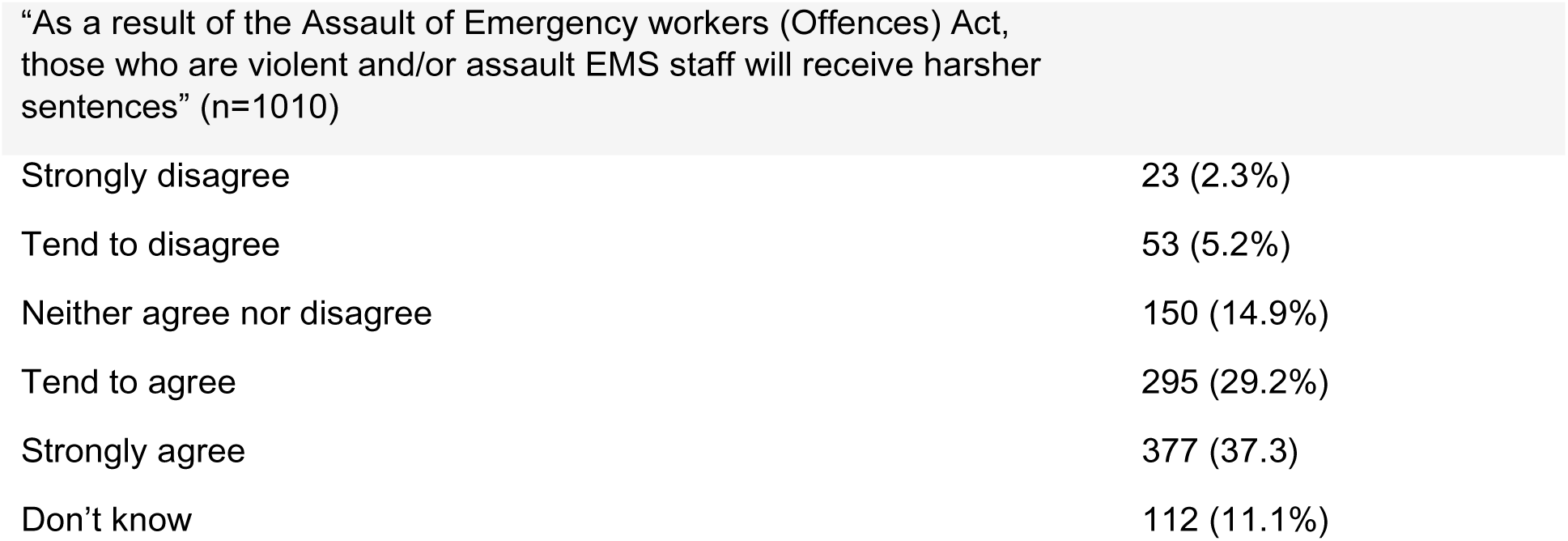
Tabulation of “To what extent do you agree or disagree with the statement?”

|  |  |
| --- | --- |
| Q13) To what extent do you agree or disagree with the statement? | Frequency (percentage) |
| Strongly disagree | 23 (2.3%) |
| Tend to disagree | 53 (5.2%) |
| Neither agree nor disagree | 150 (14.9%) |
| Tend to agree | 295 (29.2%) |
| Strongly agree | 377 (37.3) |
| Don't know | 112 (11.1%) |

53.3% of participants were not aware of any publicity and/or media campaigns launched in the last two years asking people to treat emergency workers with respect before taking this survey, whilst 46.7% were aware, table 46. Females [OR 1.321, 95% CI 1.027-1.699; p=0.030] were more aware of publicity and/or media campaigns launched in the last two years, table 47.

**Table 45:** Ordinal logistic regression of “To what extent do you agree or disagree with the following statement?”

|  |  |  |
| --- | --- | --- |
| Female | 0.949 [0.745, 1.208] (p=0.669) | 0.942 [0.737, 1.203] (p=0.631) |
| Age | 1.018 [1.011, 1.025] (p<0.001) | 1.018 [1.010, 1.026] (p<0.001) |
| C2/D/E | 0.986 [0.775, 1.255] (p=0.909) | 0.969 [0.758, 1.238] (p=0.801) |
| Welsh region (Reference category = Cardiff) |  |  |
| Mid and West Wales | 1.143 [0.756, 1.727] (p=0.528) | 0.926 [0.605, 1.418] (p=0.724) |
| North Wales | 1.166 [0.772, 1.762] (p=0.465) | 1.081 [0.711, 1.642] (p=0.716) |
| South Wales Central | 1.154 [0.662, 2.014] (p=0.613) | 1.078 [0.614, 1.891] (p=0.794) |
| South Wales East | 1.191 [0.799, 1.775] (p=0.392) | 1.173 [0.784, 1.754] (p=0.437) |
| South Wales West | 1.168 [0.757, 1.802] (p=0.483) | 1.114 [0.720, 1.724] (p=0.628) |
| ALL WORKERS (NET) | 0.724 [0.568, 0.921] (p=0.009) | 0.914 [0.700, 1.192] (p=0.507) |

Note: \* The category 'don't know' has been excluded from the model as the response is not ordered.

**Table 46:** Tabulation of “Were you are aware of any publicity and/or media campaigns that launched in the last two years (i.e. since August 2020) asking people to treat emergency workers with respect?”

| Q14) Were you are aware of any publicity and/or media campaigns that launched in the last two years (i.e. since August 2020) asking people to treat emergency workers with respect? (n=1010) | Frequency (percentage) |
| --- | --- |
| No, I wasn't | 538 (53.3%) |
| Yes, I was | 472 (46.7%) |

**Table 47:** Binary logistic regression of “Were you are aware of any publicity and/or media campaigns that launched in the last two years (i.e. since August 2020) asking people to treat emergency workers with respect?”

|  |  |  |
| --- | --- | --- |
| Female | 1.308 [1.020, 1.676] (p=0.034) | 1.321 [1.027, 1.699] (p=0.030) |
| Age | 1.005 [0.998, 1.012] (p=0.192) | 1.008 [1.000, 1.016] (p=0.051) |
| C2/D/E | 0.808 [0.631, 1.036] (p=0.093) | 0.843 [0.655, 1.085] (p=0.184) |
| Welsh region (Reference category = Cardiff) |  |  |
| Mid and West Wales | 0.843 [0.542, 1.312] (p=0.449) | 0.840 [0.535, 1.320] (p=0.450) |
| North Wales | 0.760 [0.491, 1.175] (p=0.217) | 0.759 [0.489, 1.180] (p=0.221) |
| South Wales Central | 0.758 [0.422, 1.360] (p=0.353) | 0.769 [0.426, 1.387] (p=0.382) |
| South Wales East | 0.993 [0.649, 1.520] (p=0.976) | 0.994 [0.648, 1.525] (p=0.978) |
| South Wales West | 0.807 [0.512, 1.271] (p=0.355) | 0.815 [0.516, 1.289] (p=0.382) |
| ALL WORKERS (NET) | 1.116 [0.871, 1.429] (p=0.385) | 1.212 [0.921, 1.594] (p=0.170) |

*Note:* \* The category 'don't know' has been excluded from the model as the response is not ordered.

## Discussion

We believe this is the first UK survey of public attitudes to V&A towards EMS staff. We have reported elsewhere recent efforts to tackling this issue, and how it has become a priority within Wales (UK), resulting in a wide range of policy developments and initiatives such as the Assaults on Emergency Workers (Offences) Act (2018) JESG #WithUsNotAgainstUs campaign (Rees et al 2021). Our results reflect an awareness of this issue by the public, which may or may not reflect the impact of these initiatives and may be a useful benchmark for future evaluations. Our results also reveal more scope for raising awareness, as 24.4% had never heard about V&A directed toward EMS staff.

Most participants (72.1%) had never witnessed V&A directed toward EMS staff, but worryingly,15.5% had done so less than 10 times. Younger participants were more likely to have witnessed more incidents of V&A towards EMS staff, which may be due to their presence at social settings where EMS staff may be likely to face V&A such as pubs serving alcohol. Of those who had heard about V&A towards EMS staff, 17.7% had heard via work, with workers, females and younger participants being more likely to have heard via this situation.

Most (81.3%) participants that had heard about V&A directed towards EMS staff, did not hear via a social setting (e.g. pub, sports team, etc.). Social settings present an opportunity to do target activities towards younger participants who are more likely to have hear via social settings. The most prominent situation participants reported having heard about V&A incident/s however was through the media with 81.1% of participants who had heard hearing through this method, especially older participants. Most participants strongly disagree (82.0%) with the statement, V&A can be acceptable in some cases” with a further 8.4% of participants disagreeing. Despite this, younger participants, and those with a social grade of C2/D/E were more likely to agree the statement. This may reflect their social environment where V&A may be a more accepted part of everyday life, and therefore future initiatives may wish to target attention on such groups, but it needs to be recognised that the overwhelming majority disagreed.

Limited epidemiology of the perpetrators of V&A towards EMS staff exists in the published literature. WAST (2023) however reported that offenders aged 26-35 account for the highest portion of offending (23.6%), and Friday and Saturday nights present the highest number of emergency worker assaults, accounting for 26.2% of incidents and alcohol intoxication continues to present as the largest impact factor, applying to a quarter of incidents. Evidence from other settings such as the Emergency Department have found perpetrators to be predominantly male (Nikathil et al 2018, 2017, Knott et al 2005), 52 – 81.6% of cases are associated with mental behavioural disorders (MBD) or significant mental illness, 47% of which requiring psychiatric admission (79%, involuntarily), 11% have a history of violence, 24% are repeat perpetrator (93.5% associated with a MBD), 30% involve alcohol, 17% – 42.4% involve psychoactive substance or illicit drugs (Knott et all 2005, Thomas et al 2022). In our survey most people agreed that intoxication with alcohol or drugs was a factor more likely to influence V&A towards EMS staff and that for those offenders the change in law was unlikely to deter them

WAST (2023) recently reported 1,421 assaults in the six-month period between January – June 2022 and an average of 241 per month. Our survey indicates that the public are not aware of the scale of assaults against EMS staff; for instance, only 33.2% of participants thought there were more than 500 incidents were reported to the police each year in Wales. UK/Wales based data is lacking, but across all measures, the public underestimated the scale of V&A directed towards EMS reported in the international literature, where between 67%-99% of staff report experiencing verbal abuse, 41%-80% report intimidation, 26%-69% report physical assaults,10%-17% report threats with weapons,14%-61.5% report sexual harassment and 3%-13.8% sexual assault (Bigham et al 2014, Boyle et al 2007, Savoy et al Mausz et al 2021, Newbury-Birch et al 2017, Mausz & Johnston, 2019, Suserud et al, 2002, Maguire et al., 2018).

Only 58.7% thought it was likely that EMS staff members would experience sexual assault/harassment from members of the public with women being more likely to report this. This reflects gender differences in actual sexual harassment/assault of EMS staff. Whilst both men and women EMS staff report sexual harassment, females are significantly more often affected; 24%-34.2% of females versus 6%-9.2% of males (Savoy et al 2021, Shabanikiya, et al 2021). A similar pattern is observed in other settings (Akhter et al 2019, Ansoleaga et al 2019, Chappell et al 200, 2006, Rotundo et al 2001). We have previously reported EMS staff experiences of sexual V&A which reflect international experiences, especially for women and how sociocultural constructs of misogyny and sexual V&A within EMS and its culture have not been effectively tackled (Rees et al 2023). Since conducting this study, this has received high profile media attention (BBC 2023) and the Association of Chief Ambulance Executives (AACE) and Office of the Chief Allied Health Professions Officer (CAHPO) have launched publications aimed at reducing misogyny and improving sexual safety in the ambulance service AACE (2023).

In our survey, 64.2% of participants thought it was likely that the patient being treated by EMS staff would be violent and/or aggressive towards them while they are trying to treat the patient; whilst 23.0% thought it was unlikely. Our participants underestimated the likelihood of patients, relatives, or their friends being violent and/or aggressive towards EMS staff; 20.00% however felt it was not very likely, despite having been reported to be common perpetrators (Maguire and Hunting, Bigham et al., Taylor et al., 2015; Gormley et al., 2016; Rahmani et al., 2012). People with a social grade C2/D/E were more likely to accurately think a relative/friend of the patient would be violent and/or aggressive towards EMS staff than people which may reflect actual experiences of V&A in this population?

Most participants accurately recognised the role of intoxication with alcohol and drugs as a contributory factor to V&A directed towards EMS staff. Females and older participants were more likely to think intoxication with drugs contributed to V&A directed towards EMS staff. 84.3% felt it was was likely that altered mental status following illness and/or injury make people more likely and contributes to being aggressive and/or violent towards EMS staff; while 6.5% felt it was unlikely. No characteristics were indicative of whether a participant thought an altered mental status following illness and/or injury contributed to V&A towards EMS staff.

Most of our participants (78.0%) were not aware of the Assaults on Emergency Workers (Offences) Act of 2018 which should be of concern. Most felt it was unlikely (75.2%) to deter those intoxicated with alcohol from being violent and/or aggressive towards EMS staff, and only 15.4% thought it was likely. It was somewhat reassuring that males report more awareness of this Act. Most (75.6%) thought the act was unlikely to deter those with an altered mental status following illness and/or injury and only 13.0% thought it was likely to. Views were mixed on whether other members of the public would be deterred from being violent and/or aggressive towards EMS staff, 42.9% thought it was likely and 42.6% it was unlikely.

Younger participants were also more likely to think the Act would deter people intoxicated with alcohol, drugs, people with an altered mental status following illness and/or injury and other members of the public. This is encouraging as, highlighted earlier, younger people were more likely to be perpetrators of such violence and/or aggression. This raises significant questions around the purpose of the act and may indicate the need for other interventions as well as raising awareness of legal consequences among the public. Such as, how can EMS staff be protected or protect themselves? can interventions be developed with the public which recognise the challenge of people becoming violent when they are intoxicated? and how can it be stopped? Working with offenders to understand what would have made a difference to stopping them from assaulting staff. These are very difficult issues, and a combination of approaches will likely be needed. 66.5% agreed with the statement, “as a result of the Assault on Emergency workers (Offences) Act, those who are violent and/or assault EMS staff will receive harsher sentences.” However, older participants were more likely have think offenders would receive harsher sentences because of the act which may reflect support for punitive approaches to this issue?

Public education messages based on theory are more effective than those that are not (Guttman, 2015, Tay 2011). 81.1.% of those in our study who reported hearing about V&A had done so through the media, which reflects previous literature recognising how use of mass media, including television, radio, internet, and printed materials, is still the best way to spread a message for short-term or long-term campaigns as they keep the message current, reach the largest audiences, and cost little per audience member (Wakeman, Loken, & Hornik 2010). These media campaigns should come from many directions and with a persistent and consistent message (Wakeman, Loken, & Hornik, 2010). The World Health Organization therefore recommends using media campaigns as a way to change attitudes, behaviors, and social norms with regard to violence (Vivolo, Matjasko, & Massetti, 2011). 46.7% of participants were aware of publicity and/or media campaigns launched since August 2020 asking people to treat emergency workers with respect. Females were more aware of publicity and/or media campaigns. Efficacy of communication campaigns varies greatly (Cho & Salmon, 2007) and they can have a wider range of impacts, including positive (e.g., Devlin, Eadie, Stead, & Evans, 2007), no effects at all (Foxcroft et al 1997), and negative effects (Bushman et al 2012). Faced with such disparate results, it is difficult to anticipate when and, for whom, anti-violence campaigns will be effective, ineffective, or even counterproductive (Merrell et al 2008).

Previous research on violent incidents has found that communication campaigns do not seem to achieve their goal of reducing the problem (Cárdaba et al 2016). Messages that contain prohibitions or seek to change behaviour through fear appeals tend to be ineffective or even counter-productive for reducing violent attitudes and aggressive behaviour (Bushman & Stack 1996, Roskos-Ewoldsen, Yu, & Rhodes, 2004, Cárdaba et al 2016). The Wales anti-violence campaign asked the public to work with us not against, us and treat staff with respect whist highlighted the consequence of V&A on them. This campaign may however have introduced an element of fear, highlighting that V&A on EMS staff was not acceptable and could result in a prison sentence.

Reactance is deemed the state of psychological activation and resistance that arises when our freedom is limited or threatened: the most direct consequence of this state is a tendency to resist everything that could be considered as a threat to one’s personal liberty (Brehm & Brehm 2013). Such messages are often perceived as threatening and can generate reactance from the intended audience (Byrne, Linz, & Potter, 2009; Kim, Levine, & Allen, 2014) resulting in campaigns failing, and even backfiring resulting in favourable attitudes in the target population (Nyhan, Reifler, Richey, and Freed 2014). The public appear to support the notion that V&A towards EMS staff is not acceptable and we have identified groups and settings were this message may be amplified, such as young people, men, those in social grade of C2/D/E and through social setting. Targeting such populations and settings where this issue may be more prevalent seems sensible, but if the aim is to prevent V&A towards EMS staff, it is currently unknown if there are any wider costs to the efforts of communication campaigns, policy and legislation and if they are indeed effective or harmful?

Eburn, and Townsend, (2018) provide sophisticated commentary in relation to jail sentences for perpetrators of V&A on EMS staff which may be relevant to wider initiatives above. They highlight that in order to protect EMS staff, the offending needs to be affected before it happens. Harsh sentencing for perpetrators aimed at protecting emergency workers from future violence they suggest is akin to delivering a placebo (“A … procedure prescribed for the psychological benefit to the patient rather than for any physiological effect” or “A measure designed merely to humour or placate someone”) rather than effective or meaningful protection. Eburn, and Townsend, (2018) do however acknowledge that punitive sentences for people who assault paramedics may be called for in some circumstances, that it is not okay to intentionally assault paramedics, and that’s why it’s illegal, as it should be.

It may be argued that legislation and high-profile communication may act as a deterrent for V&A on EMS staff, but this may rely on accurately targeting potential perpetrators which may be challenging, and as discussed earlier, could have the opposite effect. Eburn, and Townsend, (2018) highlight the political dimension to this issue as politicians can win many more votes by being ‘tough on crime’. Even when there is awareness in these groups, the preventative capacity of such initiatives may be limited, as most perpetrators may be intoxicated with drugs and alcohol or incapacitated through illness or injury at the time of the V&A. Our study suggests the public are also aware of the limitations of such initiatives due to the impact of intoxication. Eburn, and Townsend, (2018) once again point to the potential costs of such punitive approaches through its potential to reduce trust of EMS in these vulnerable patient groups and communities. Eburn, and Townsend (2018) urges caution over governments and others and appearing to act to placate to those calling for something to be done, but rather call for tackling the causes of V&A by funding education, provision of mental health services, drug and alcohol rehabilitation.

## Limitations

This study is limited as it did not involve a probabilistic sample and does not intend to establish causal links, we do however report associations among participant characteristics. Members of our team have used similar methods through YouGov in UK surveys of attitudes to Cardiopulmonary Resuscitation and Defibrillator Use (Hawkes et al 2017), which continue to be valued by the ambulance services community. By using a prespecified sample size, our approach was able to sample this appropriate population quickly and efficiently, which would otherwise have been extremely challenging and costly by other survey methods.

## Conclusion

Understanding public attitudes towards protecting EMS staff from violence and aggression is important, to consider the societal impact of policy, legislation and initiatives such as information campaigns. Our study has revealed that the public have good awareness this issue and appear to support the notion that V&A towards EMS staff is not acceptable but underestimate the actual scale of the problem. We have identified groups and settings where this message may be amplified, such as young people, men, those in social grade of C2/D/E and through social settings where V&A may be encountered more often. We also found that females had great awareness and perception of sexual harassment and assault which reflects the actual experiences of female EMS staff who are more likely to encounter this form of V&A. We, therefore, recommend future campaigns targeted towards men, more research in to why and what interventions could help and research to quantify the actual scale of sexual assault and harassment for ambulance staff from the public and internally.

Participants were however less enthusiastic about the likelihood of the effectiveness of legislation in deterring those with drugs and alcohol or altered mental status following illness and/or injury who represent the likely perpetrators of V&A towards EMS staff. It is therefore currently unknown whether such legislation, policy initiatives and information campaigns will have any impact on the actual problem. We recommend UK wide survey using this methodology as a benchmark. We also recommend further research needs to focus on the epidemiology of perpetrators and development of evidence-based interventions to protect EMS Staff from Aggression and Violence in Conflict Encounters.

## Data Availability

All data produced in the present work are contained in the manuscript

## Acknowledgments

YouGov conducted the survey on behalf of the PEACE research collaborators and Welsh Ambulance Services NHS Trust (WAST) through the Pre Hospital Emergency Research Unit (PERU). We would like to thank WAST & PERU for sponsoring and funding the study, along with collaborators from Kings College London Swansea Trials Unit (STU) who provided statistical analysis support.

